# Symptom experience before vs. after confirmed SARS-CoV-2 infection: a population and case control study using prospectively recorded symptom data

**DOI:** 10.1101/2023.08.30.23294821

**Authors:** Carole H. Sudre, Michela Antonelli, Nathan J Cheetham, Erika Molteni, Liane S Canas, Vicky Bowyer, Ben Murray, Khaled Rjoob, Marc Modat, Joan Capdevila Pujol, Christina Hu, Jonathan Wolf, Tim D Spector, Alexander Hammers, Claire J Steves, Sebastien Ourselin, Emma L Duncan

**Affiliations:** School of Biomedical Engineering & Imaging Sciences, King’s College London, London, UK; MRC Unit for Lifelong Health and Ageing at UCL, Department of Population Science and Experimental Medicine, University College London, London, UK; Centre for Medical Image Computing, Department of Computer Science, University College London, London, UK; Department of Twin Research and Genetic Epidemiology, King’s College London, London, UK; ZOE Limited London, UK; Guy’s and St Thomas’ PET Centre, Guy’s and St Thomas’ NHS Foundation trust, London, UK; Department of Ageing and Health, Guy’s and St Thomas’ NHS Foundation trust, London, UK; Department of Endocrinology, Guy’s and St Thomas’ NHS Foundation Trust, London, UK

**Keywords:** COVID-19 symptoms, long COVID, post-COVID

## Abstract

**Background:** Some individuals experience prolonged illness after acute COVID-19. We assessed whether pre-infection symptoms affected post-COVID illness duration.

**Methods:** Survival analysis was performed in adults (n=23,452) with community-managed SARC-CoV-2 infection prospectively self-logging data through the ZOE COVID Symptom Study app, at least weekly, from 8 weeks before to 12 weeks after COVID-19 onset, conditioned on presence vs. absence of baseline symptoms (4-8 weeks before COVID-19). A case-control study was performed in 1350 individuals with long illness (≥8 weeks, 906 [67.1%] with illness ≥12 weeks), matched 1:1 (for age, sex, body mass index, testing week, prior infection, vaccination, smoking, index of multiple deprivation) with 1350 individuals with short illness (<4 weeks). Baseline symptoms were compared between the two groups; and against post-COVID symptoms.

**Findings:** Individuals reporting baseline symptoms had longer post-COVID symptom duration (from 10 to 15 days) with baseline fatigue nearly doubling duration. Two-thirds (910 of 1350 [67.4%]) of individuals with long illness were asymptomatic beforehand. However, 440 (32.6%) had baseline symptoms, vs. 255 (18.9%) of 1350 individuals with short illness (p<0.0001). Baseline symptoms increased the odds ratio for long illness (2.14 [CI: 1.78; 2.57]). Prior comorbidities were more common in individuals with long vs. short illness. In individuals with long illness, baseline symptomatic (vs. asymptomatic) individuals were more likely to be female, younger, and have prior comorbidities; and baseline and post-acute symptoms and symptom burden correlated strongly.

**Interpretation:** Individuals experiencing symptoms before COVID-19 have longer illness duration and increased odds of long illness. However, many individuals with long illness are well before SARS-CoV-2 infection.

## Introduction

SARS-CoV-2 has infected more than half a billion individuals to date. Individuals of older age, male sex, and with prior comorbidities have poorer outcomes after acute infection, including higher rates of hospitalization and mortality [1]. many individuals hospitalized for COVID-19 experience protracted convalescence [2,3] particularly individuals requiring ventilatory support; a longitudinal UK study found the majority (71%) were not fully recovered six months post-discharge [4]. However, many community-managed individuals also report protracted post-acute illness. An early community-based study found 13.3% experienced illness beyond 4 weeks, and 2.8% beyond 12 weeks, with longer duration associated with female sex, older age, more severe acute illness, and prior comorbidities [3]. In the UK, ongoing symptomatic COVID-19 (OCS) and the Post COVID-19 syndrome (PCS) are defined as otherwise-unexplained symptoms and signs for 4-12 weeks (OSC), or more than 12 weeks (PCS), after an acute illness attributable to SARS-CoV-2 infection [5] with some variation internationally in terminology and symptom duration (e.g., 8 vs. 12-week threshold) [6]).

PCS prevalence estimates vary substantially. A recent meta-analysis [7] highlighted the heterogeneity of published studies (*I*^2^ = 100%) with widely differing prevalence estimates (9%-81%), varying globally, regionally, and by hospitalization status. PCS prevalence estimates derive from predominantly hospitalized cohorts. In March 2023 the UK Office of National Statistics estimated 1.9 million citizens (2.9% of the population) had self-reported long COVID (defined as symptoms for more than four weeks after either test-positive or suspected SARS-CoV-2 infection)[8]. An earlier UK study reported prevalence of PCS of 1.2%-4.8% in test-positive individuals, considering only symptoms that limited day-to-day functioning [9]. Neither study included a control group. In contrast, a large UK primary-care study comparing community-managed adults with confirmed SARS-CoV-2 infection to a matched contemporaneous cohort reported symptom prevalence (here, at least one symptom) at 12 weeks of 5.4% in infected vs. 4.3% in uninfected individuals [10].

Ongoing and/or post-acute symptomatology after acute infection has been reported after many infections, including bacteria (e.g., Borrelia) viruses (e.g., Epstein-Barr Virus) and parasites (e.g., Giardia), which share many characteristics including fatigue, exertional intolerance, and neurocognitive symptoms (‘brain fog’) (recently comprehensively reviewed [11]). Post-acute infection syndromes are more common in females and younger individuals, though a relationship with initial illness severity is less clear [11]. Few studies assess pre-morbid risk factors for post-acute syndromes prospectively [12] despite possible influence of premorbid conditions on post-acute illness symptomatology [13]. Postulated non-exclusive mechanisms include remnant infection, autoimmunity induction, and/or maladaptive tissue repair; however, for most affected individuals, definitive pathophysiology is unclear despite extensive investigations. Whether similar processes underpin PCS, and whether common to all PCS individuals, is unclear [14].

The ZOE COVID Symptom Study began in March 2020 with participating adults logging their health data contemporaneously across the pandemic. Thus, symptoms could be assessed prospectively and longitudinally in individuals subsequently contracting SARS-CoV-2, and irrespective of ultimate illness profile.

We hypothesised that symptoms and comorbidities before SARS-CoV-2 infection might contribute to post-acute symptomatology, including illness duration. We assessed:

A) symptoms reported before COVID-19, in individuals subsequently experiencing long vs. short illness;
B) symptom correlation before and after SARS-CoV-2 infection; and
C) whether prior symptoms and comorbidities affect post-COVID illness profile.

## Methods

### Cohort

The ZOE COVID Symptom Study launched in the UK on 24 March 2020 as a collaboration between ZOE Ltd, King’s College London, Massachusetts General Hospital, Lund University and Uppsala University (ethics approval: KCL ethics committee REMAS no. 18210, review reference LRS-19/20– 18210, with all individuals providing informed consent for use of their data in COVID-19 research at registration). After initial logging of baseline demographics including comorbidities, (**Suppl. Table S1**) participants were prompted daily to report any symptoms (direct questions and free text (**Suppl. Table S1**)), SARS-CoV-2 testing and results, and vaccination(s) using a phone-based app. Data collection expanded on 4 November 2020 to include more direct symptom questions. The cohort was surveyed regarding pre-pandemic mental health diagnoses in February-April 2021 [15] (**Suppl. Table S2**). The current dataset was cut on 30 May 2022 with symptom assessment altering the following day.

As previously [3], COVID-19 was defined as symptoms associated with SARS-CoV-2 infection (**Suppl. Table S1**) commencing between 14 days before and 7 days after a self-reported positive polymerase chain reaction (PCR) or lateral flow antigen test (LFAT). For individuals with multiple positive tests, subsequent illness profiles were defined for tests spaced >90 days apart. If ≤90 days apart, illness profile was defined according to the first positive test. Illness duration was calculated from first symptomatic day until return to asymptomatic (i.e., logging as healthy) [3,16]. Consideration was given to possible right censoring in duration calculation (ongoing illness at final data censoring; logging discontinuation while still symptomatic). To calculate illness duration attributable to acute SARS-CoV-2 infection, individuals were required to log as healthy for at least one week immediately before COVID-19 commencement [3,16]. If symptoms were again logged within one week of a healthy report, illness was considered ongoing, thus allowing for illness fluctuation [3,16].

The baseline period was defined as 4 to 8 weeks before, and the post-COVID period as 8-12 weeks after, COVID-19 onset. To ensure consistent assessment in the entire cohort for the necessary 20 weeks, data were constrained to individuals in whom COVID-19 commenced between 30 December 2020 (8 weeks after 4 November 2020, date of symptom question expansion) and 2 March 2022 (12 weeks before 1 June 2022, date of symptom assessment alteration). Symptom burden was calculated as number of individual symptoms reported at least once during the defined period of assessment. Self-reported mental health diagnoses were considered overall, and for disorders that can include psychosis (Suppl. Table S2).

Short illness was defined as <4 weeks and long illness as ≥8 weeks.

Inclusion criteria were:

a) self-reporting UK adults presenting with PCR- or LFAT-confirmed COVID-19, between 30 December 2020 and 2 March 2022.
b) logging at least once weekly, from ≥8 weeks before until ≥12 weeks after COVID-19 commencement.
c) logging as healthy in the week before COVID-19 commencement.
d) co-morbidity and demographic data logged at registration, with subsequent participation in the mental health survey.

In addition to non-adherence to inclusion criteria, individuals vaccinated within either baseline or post-COVID periods or one week before these periods were excluded, given symptom overlap between vaccination side-effects and COVID-19 [17].

For remaining individuals, a Cox model was performed, adjusting for demographic criteria including age, sex, body mass index (BMI), week of test, number of test-positive SARS-CoV-2 infections, smoking status, index of multiple deprivation (IMD), and number of vaccinations, evaluating the effect of any symptom present at baseline on median duration overall, and for each symptom individually.

A matched case-control analysis was then performed. Individuals with long illness (≥8 weeks) were selected first. Individuals with short illness (<4 weeks) were then selected, matched 1:1 per previously listed demographic criteria using the Hungarian algorithm [18], minimizing the Euclidean distance cost and ensuring equal weighting across all characteristics (normalising baseline variables before matching); without replacement for controls.

Data were compared across groups using the McNemar test for counts and Wilcoxon signed rank tests for continuous variables.

Using conditional logistic regression models, we assessed the odds of long illness duration according to baseline symptom presentation (considered overall [i.e., any symptoms at baseline] and for each individual symptom) using three levels of adjustment for covariates:

1. Model 1 – no adjustment
2. Model 2: As for Model 1, with additional adjustment for presence of any comorbidity logged at registration (allergic rhinitis [hay fever], cancer, diabetes, kidney disease, heart disease, lung disease, asthma)
3. Model 3: As for Model 2, with additional adjustment for prior mental health diagnosis.

To investigate relationships between baseline and post-COVID symptoms, we assessed the odds of experiencing each individual symptom in the post-COVID period according to its presence at baseline, separately in individuals with long and with short illness, using logistic regression models with the three levels of adjustment detailed above, and adjusting for previously listed demographic variables. We further investigated possible sex-based difference in reporting at baseline and in the post-COVID period according to duration group given previous evidence of sex differences in post-acute infection syndromes [11],and OSC/PCS [8]. For symptoms with a severity scale (fatigue and dyspnoea), we compared severity during baseline and post-COVID periods in individuals reporting these symptoms at baseline; we also considered dyspnoea specifically in individuals with prior asthma/lung disease. Seasonal effects on symptom reporting were assessed for each individual symptom separately for long and short illness groups, using summertime (May-September) as reference. As some co-morbidities exhibit symptom seasonality (e.g., allergic rhinitis) adjustment for previously listed demographic variables and Model 3 variables was applied.

For individuals with long illness, we investigated demographic differences according to baseline symptomatology (≥1 symptom at baseline) using Chi-squared test for categorical and Mann Whitney U-Test for continuous variables. We investigated the relationship between baseline and post-COVID symptom burden using linear regression, adjusting for demographic variables.

For baseline comorbidities that differed between individuals with long vs. short illness (Table 1), we assessed whether presence of any of these comorbidities affected baseline symptom presentation and overall post-COVID symptom burden, using Chi-squared and Mann-Whitney U tests respectively. We investigated odds for individual baseline and post-COVID symptoms in individuals with long illness according to these comorbidities, using a logistic regression model adjusted for demographic variables.

**Table 1:**
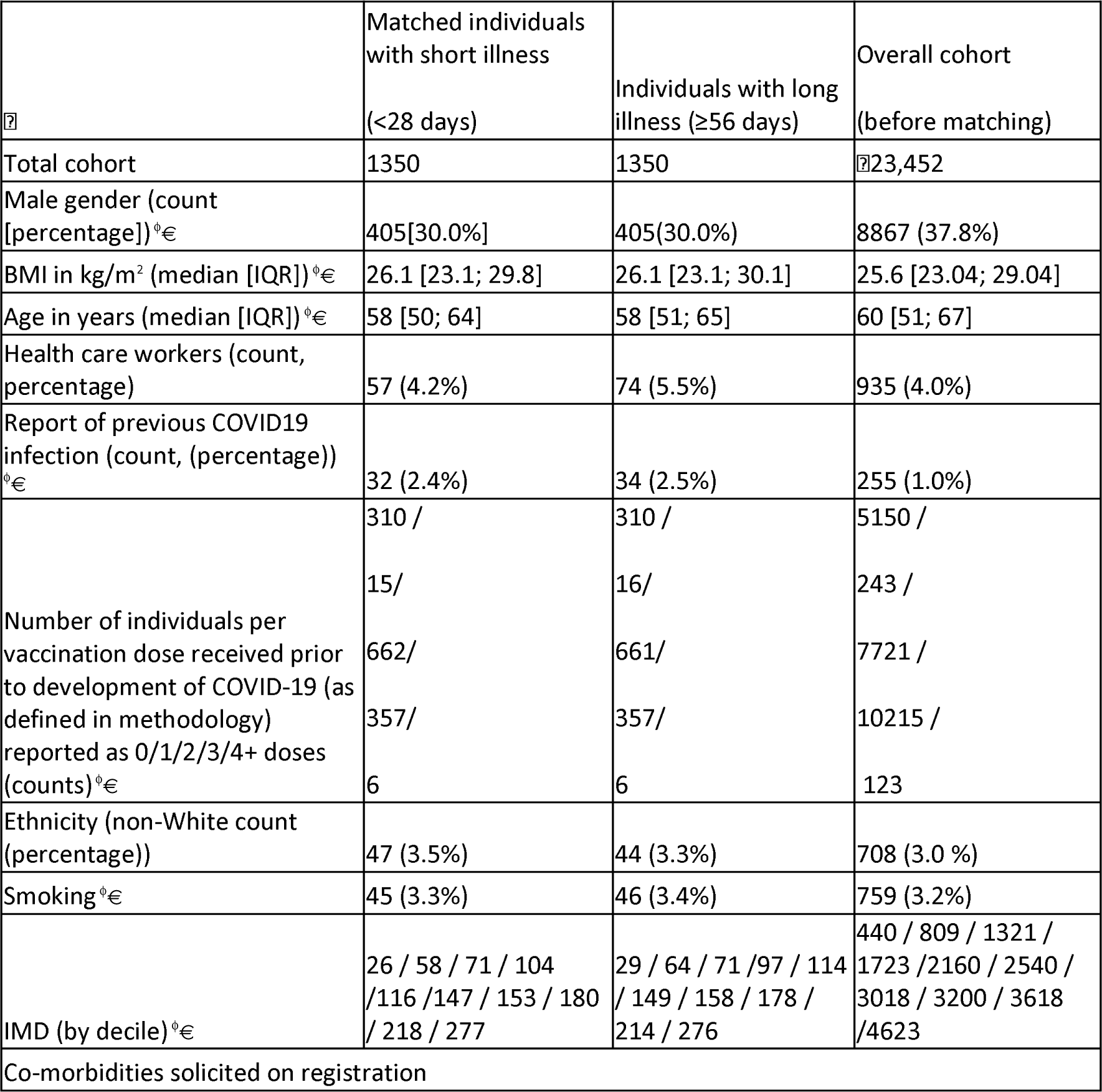
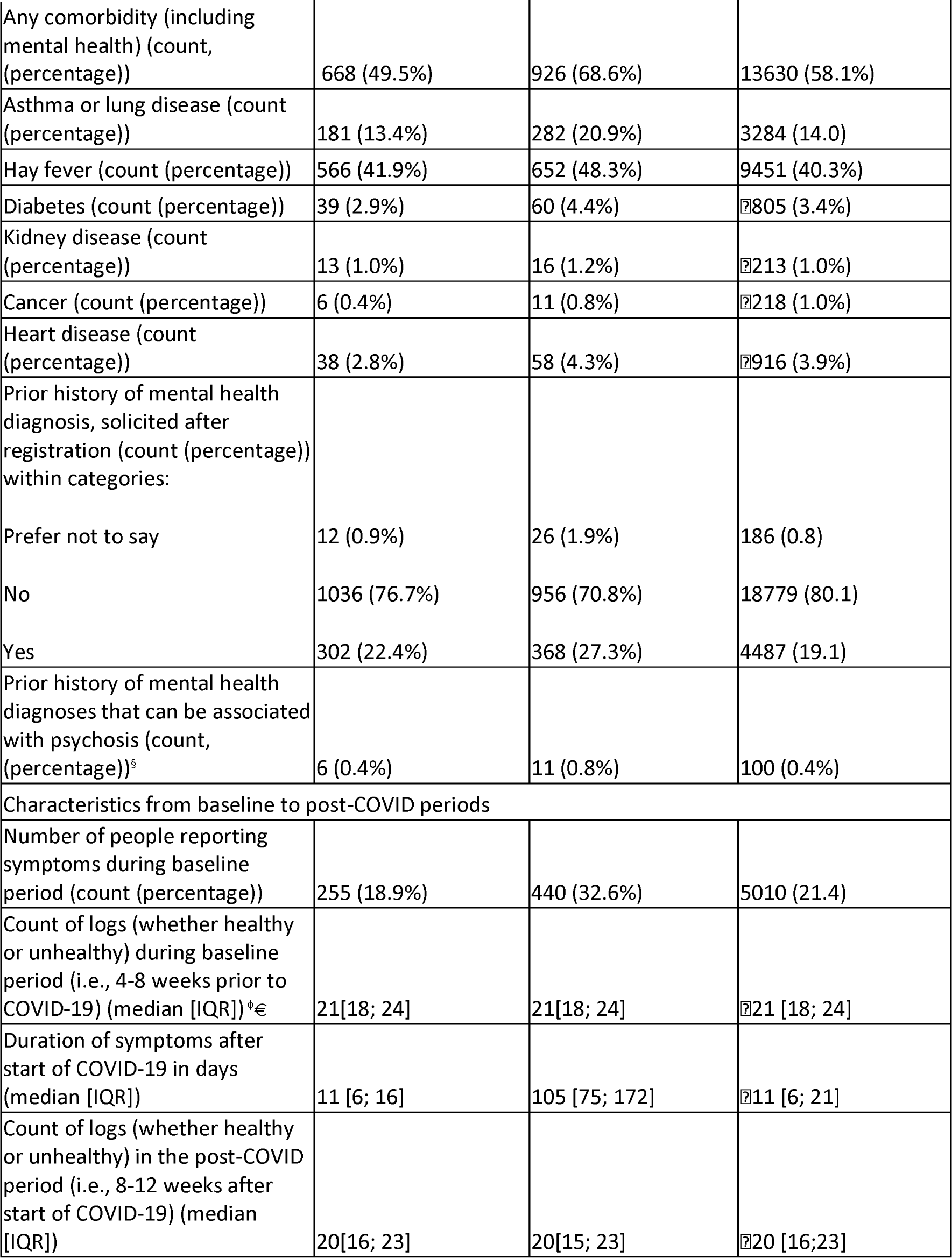
Demographic data for study participants. ⍰ indicates that the variable was used in the matching process. Continuous variables were compared using the paired Wilcoxon test; categorical variables were compared using the chi-squared test. * indicates significant difference between the two groups. § included here: self-reported categories of: mania, hypomania, bipolar, manic depression, schizophrenia, psychosis, psychotic illness. Smoking refers to currently smoking. IMD (index of multiple deprivation) is ordered from 1 (most deprived) to 10 (least deprived). Median illness duration was calculated <u>without</u> considering the possibility of right-censoring at 12 weeks in the group with long illness duration. Right censoring occurred for 338 individuals who were still logging as unhealthy at the time of study cut-off and for 166 individuals who stopped logging while still unhealthy.

We repeated the conditional logistic regression analysis and other analyses for individuals with long illness, with minimal logging frequency of at least fortnightly (vs. weekly), thus allowing for inclusion of less assiduous loggers and longer periodicity of symptom fluctuation.

False discovery rate adjustment using the Benjamini-Hochberg procedure was applied across all tested symptoms, in each analysis.

## Results

Figure 1 shows participant selection. **Table 1** presents the cohort before and after matching. Compared to the overall cohort of test-positive individuals, the selected cohort of regular loggers was older, with slightly more comorbidities. Censoring on mental health survey participation did not affect the cohort greatly except for sex (females more likely to participate); other characteristics were stable (data not shown).

**Figure 1:**
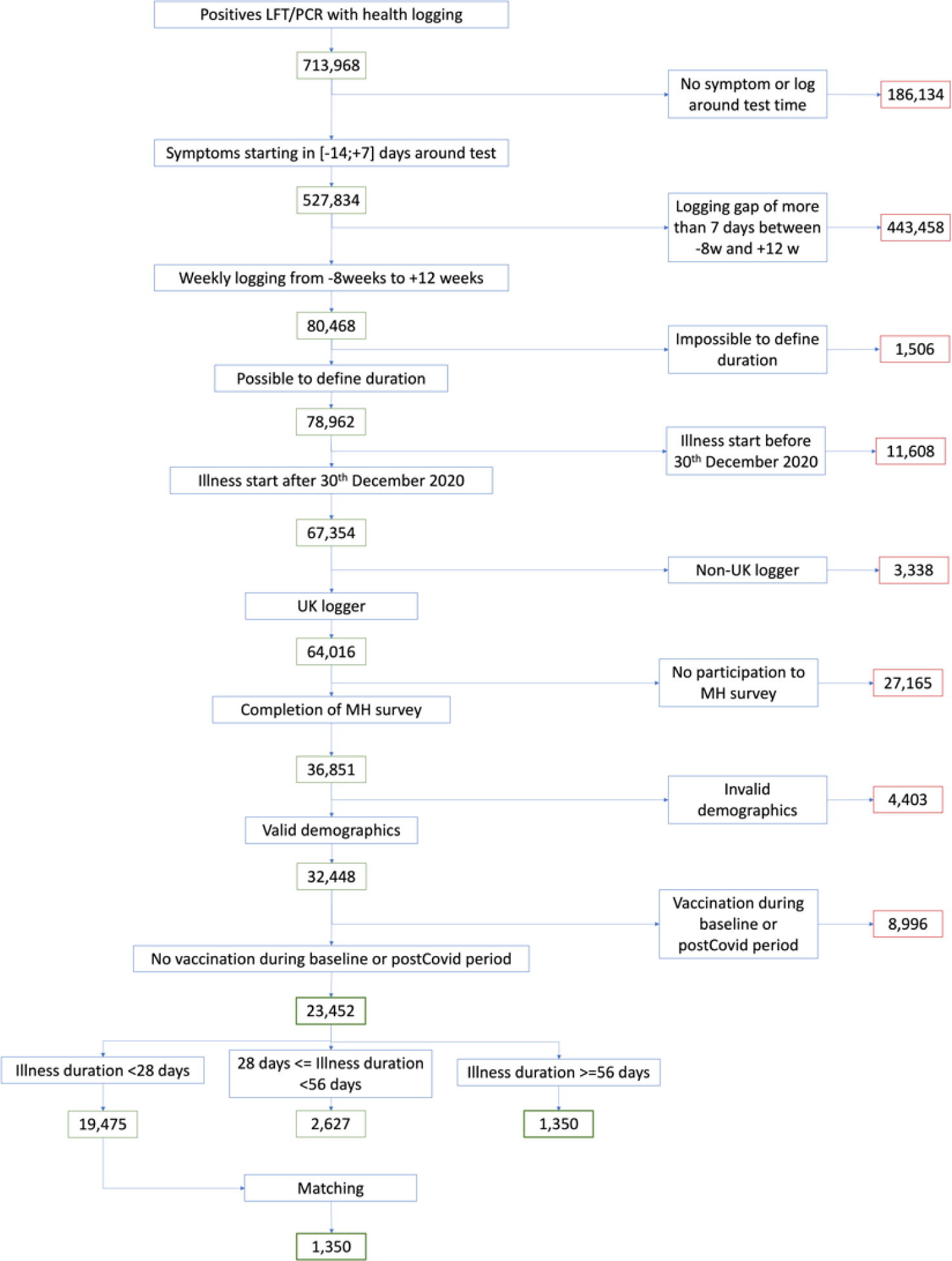
Flowchart for Study Participant Selection. LFT – lateral flow test. PCR – polymerase chain reaction test. MH – mental health. Weekly logging – at least one health report logged weekly from 8 weeks before 12 weeks after commencement of COVID-19. Invalid demographics: BMI < 15 or BMI > 55; age >100 years or age < 18 years, no possibility to extract index of multiple deprivation.

The majority (906 of 1350, 67.1%) individuals with long illness had illness duration beyond 12 weeks.

### Impact of baseline symptoms on median illness duration

Reporting of any symptom at baseline increased median illness duration (from 10 [IQR: 9;12] to 15 [IQR: 13;16] days). Right censoring (unfinished illness) was identified for some long illness individuals due to data censorship (n=338) and logging interruption while unhealthy (n=166).

Considering individual symptoms, baseline reporting of fatigue, headache, sneezing, sore throat, and rhinorrhea increased median illness duration by 9, 7, 5, 5, and 4 days respectively.

## Matched case-control cohort analysis

### Comorbidities

Considering the matched cohort (n=2700 individuals), 463 individuals reported lung disease and/or asthma: 310 (67.0%) reported both, 115 (24.8%) only asthma, and 38 (8.2%) only lung disease. Thus ‘lung disease’ and ‘asthma’ categories were jointly considered. In contrast, 1326 individuals reported allergic rhinitis and/or asthma: 317 (23.9%) reported both, 910 (67.9%) only allergic rhinitis, and 108 (8.1%) only asthma; thus, allergic rhinitis was considered separately.

Individuals with long (vs. short) illness were more likely to report co-morbidities of allergic rhinitis (p<0.001), asthma/lung disease (p<0.001), heart disease (p=0.044], diabetes (p=0.037), and/or a prior mental health diagnosis (p=0.003).

### Baseline symptoms in individuals with short vs. long illness

Individuals with long (vs. short) illness were more likely to report baseline symptoms (440 (32.6%) vs. 255 (16.8%), p-value <0.0001) (**Table 1**). However, over two-thirds (67.4%) of individuals with long illness were asymptomatic before COVID-19.

Logging frequency during the baseline period did not differ between individuals with short vs. long illness, and only marginally) in the post-COVID period (20 vs. 21 logs for individuals with long vs. short illness).

Considering individual symptoms at baseline (Figure 2), the five commonest symptoms were the same regardless of ultimate illness duration but were more prevalent in individuals with subsequent long (vs. short) illness: headache (18.9% vs. 9.4%), fatigue (13.0% vs. 5.6%), sore throat (15.0% vs. 8.8%), rhinorrhea (14.3% vs. 8.5%), and sneezing (11.9% vs.6.8%) [descriptive data only].

**Figure 2:**
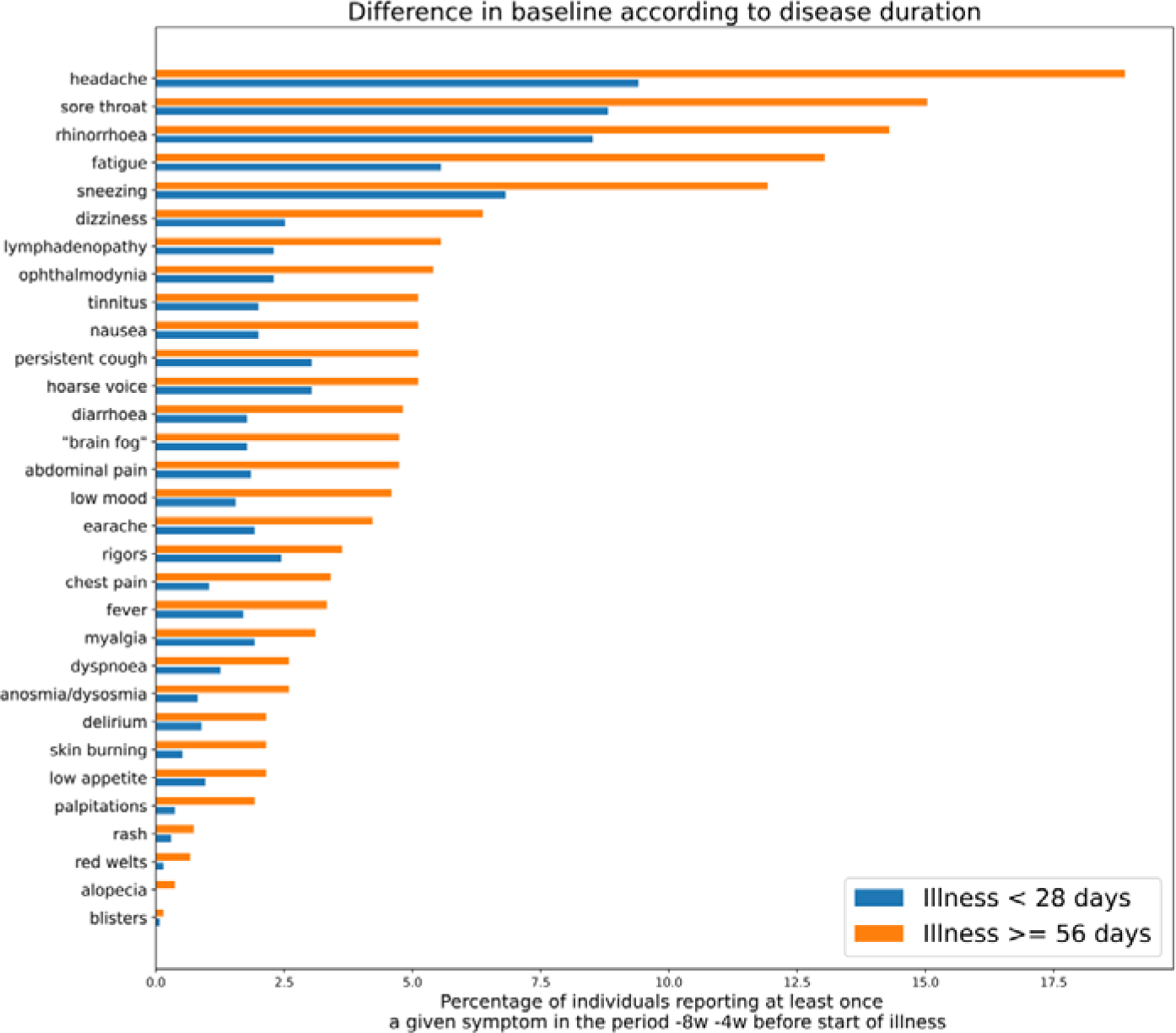
Symptom prevalence during the baseline period in individuals with long illness vs. short illness (descriptive data only, unadjusted for comorbidities, week of testing, prior infection, vaccination status, smoking or index of multiple deprivation).

### Baseline symptoms associated with increased odds of long illness

Adjusting for demographic variables, the odds ratio for long illness in symptomatic (vs. asymptomatic) baseline status was 2.14 [CI:1.78; 2.57].

Considered per-symptom, with identical covariates: reporting of almost any individual symptom at baseline increased the odds of long illness (Figure 3). However, no evidence of association was seen between long illness and baseline cutaneous symptoms (red welts, ‘blisters’, alopecia), and, after adjustment for prior comorbidities (Model 2) and prior mental health diagnoses (Model 3), anorexia (‘low appetite’).

**Figure 3:**
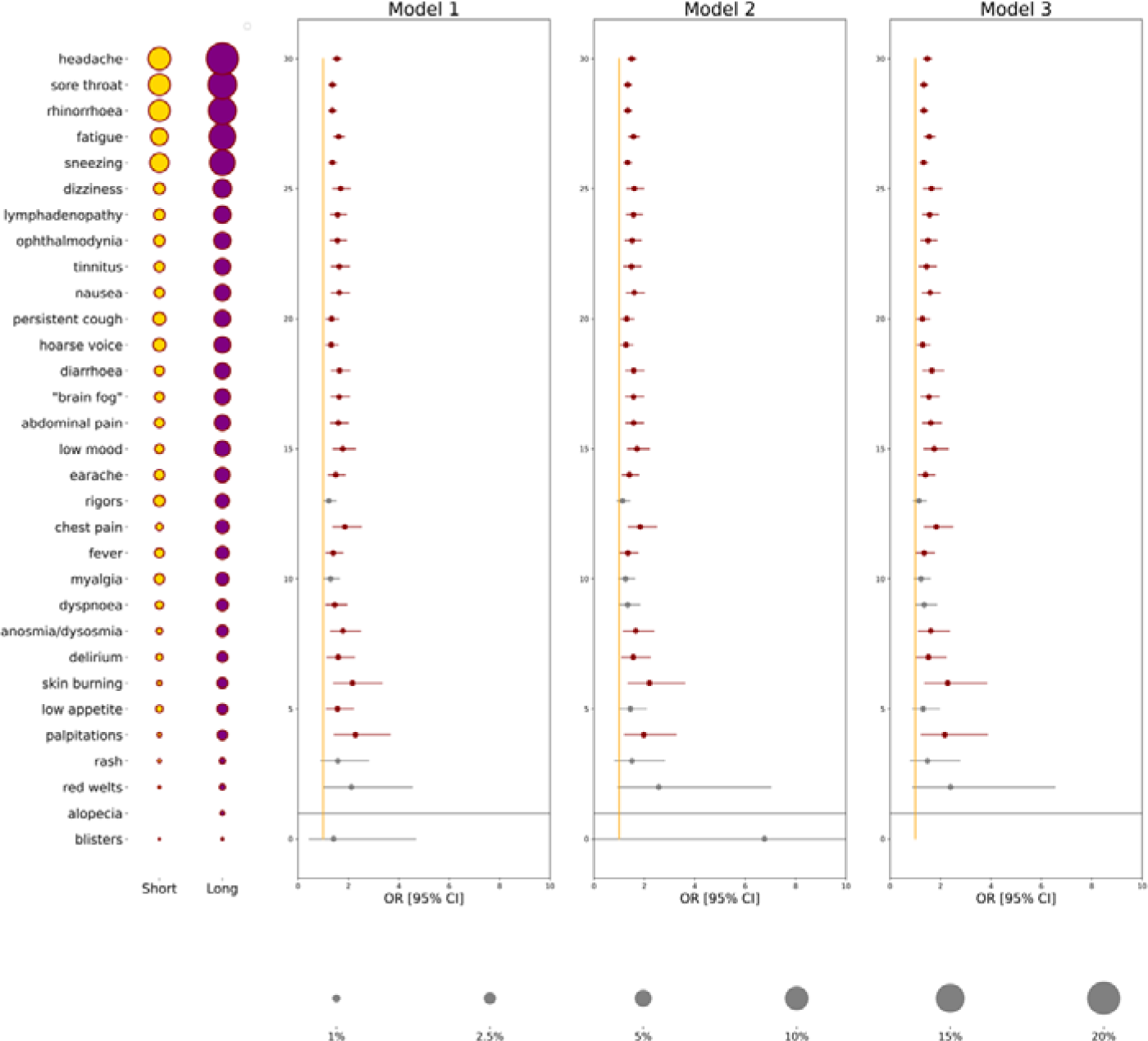
Odds ratios from conditional logistic regression of individual symptoms for long illness vs. short illness (reference group), according to baseline symptom reporting. Model 1: no adjustment; Model 2: additionally adjusted for any comorbidity reported at registration; Model 3: additionally adjusted for prior mental health diagnosis. Circle size represents baseline symptom prevalence in individuals with short (gold) vs. long (purple) illness duration; circle scale is shown at the bottom in grey. Odds ratios are shown as dots with 95% confidence intervals as lines: results in red are significant after adjustment for multiple comparisons.

### Symptom concordance over time

Figure 4 compares symptoms during **baseline or post-COVID periods,** in individuals with long or short illness (categories: stayed absent/appeared/disappeared/stayed present). Importantly, our inclusion criteria meant individuals with long illness had at least one symptom during the post-COVID period, whereas individuals with short illness had returned to asymptomatic for at least one week within four weeks of developing COVID-19 (though they might subsequently report symptoms again).

**Figure 4:**
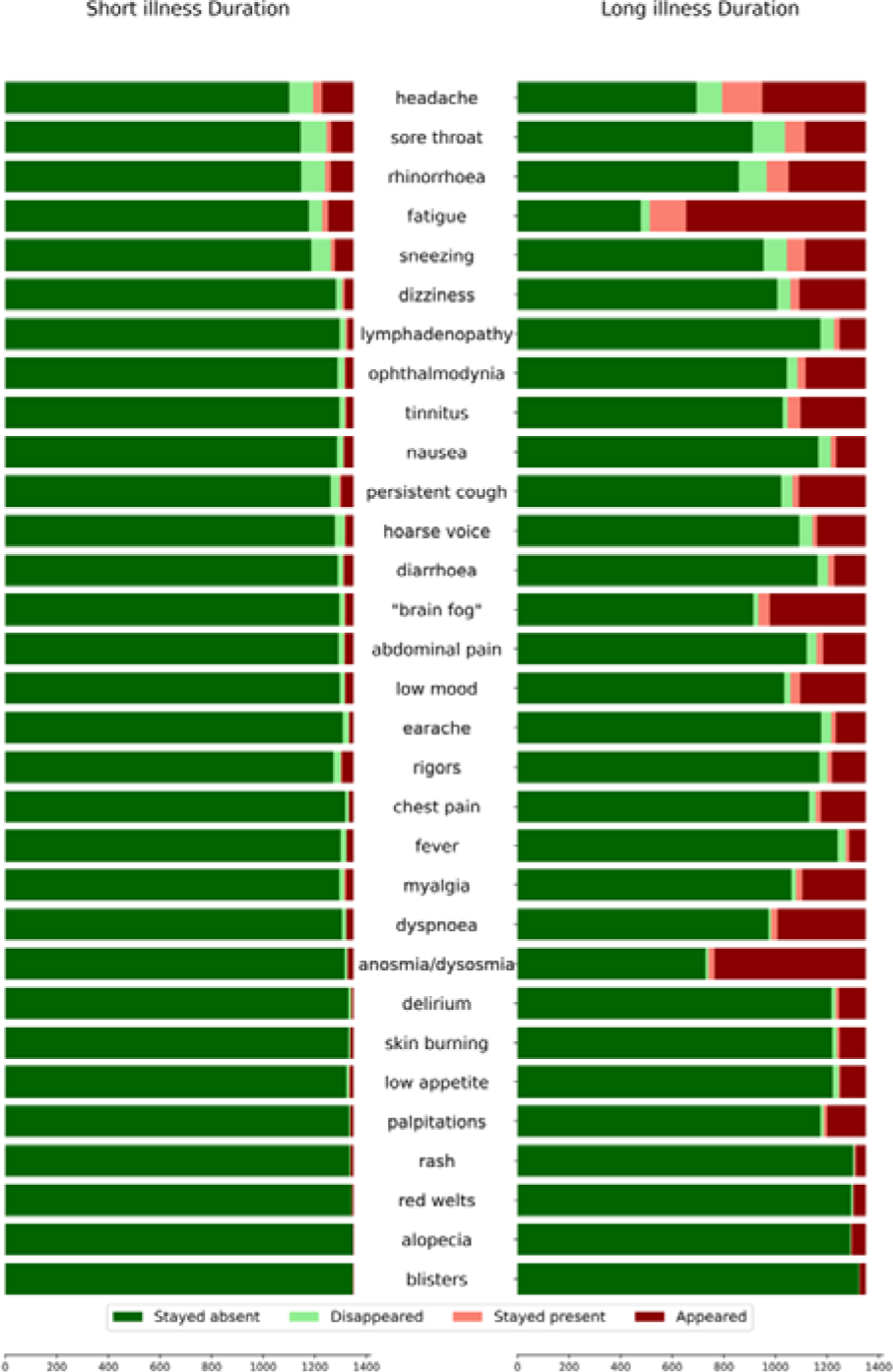
Concordance of symptoms between baseline and post-COVID periods, in individuals with short (left image) vs. long (right image) illness (n=1350 in each group).

In individuals with long illness, individual symptoms were more likely in the post-COVID period if present at baseline, with exceptions of some cutaneous manifestations (Figure 5). Adjusting for baseline comorbidities (Model 2) and prior mental health diagnosis (Model 3) made minimal difference.

**Figure 5:**
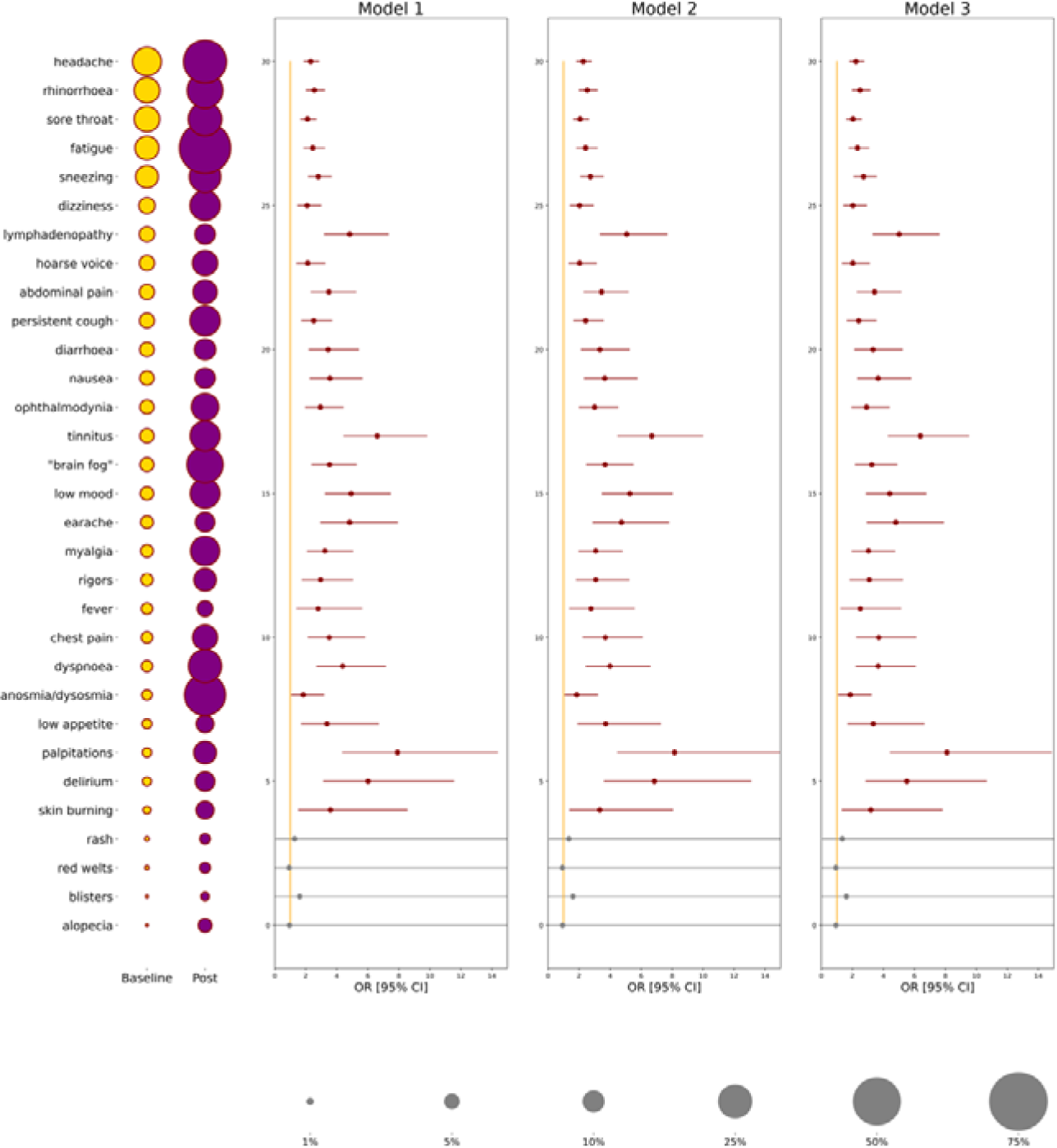
Odds ratios of symptom concordance (i.e., present in the post-COVID period, if reported at baseline [reference period]) in individuals with long illness. Model 1: adjusted for age, sex, BMI, vaccination number, prior infection, week of testing, smoking and index of multiple deprivation; Model 2: additionally adjusted for comorbidities reported at registration; Model 3: additionally adjusted for prior mental health diagnosis. Circle size refers to symptom prevalence during baseline (gold) and post-COVID (purple) periods; scale is shown at bottom of figure. Symptoms are ordered by decreasing prevalence at baseline. Odds ratios are shown as dots with CI (lines); results in red are significant after adjustment for multiple comparisons.

For comparison, and as expected given our inclusion criteria, fewer symptoms were reported in the post-COVID period in individuals with short illness, considered overall **(Suppl. Fig S1)** and by baseline individual symptom **(Suppl. Fig S2)**. Symptoms more likely to be present during baseline and post-COVID periods in this group included several upper respiratory symptoms (tinnitus, sore throat, rhinorrhoea, sneezing, hoarse voice) and some systemic symptoms (headache, lymphadenopathy, dizziness, myalgia, rigors, and fatigue). Adjusting for baseline comorbidities (Model 2) reduced significance for the rarest symptoms; adjusting further for prior mental health diagnosis (Model 3) did not alter these results substantially.

### Symptom reporting by sex

Symptom prevalence during both baseline and post-COVID periods varied numerically by sex, whether illness was of long or short duration. Most symptoms were more commonly reported by females than males **(descriptive data:** Figure 6 **and Suppl. Figure S1; statistical comparisons: Suppl. Table S3 (baseline symptoms) and Suppl. Table S4 (post-COVID symptoms**). Sex differences in symptom prevalences appeared least for post-COVID symptoms in individuals with long illness (Figure 6, lower right panel), and for baseline symptoms in individuals with short illness (Figure 6, upper left panel). These findings should be interpreted descriptively as we did not formally test for an interaction between symptoms and sex.

**Figure 6:**
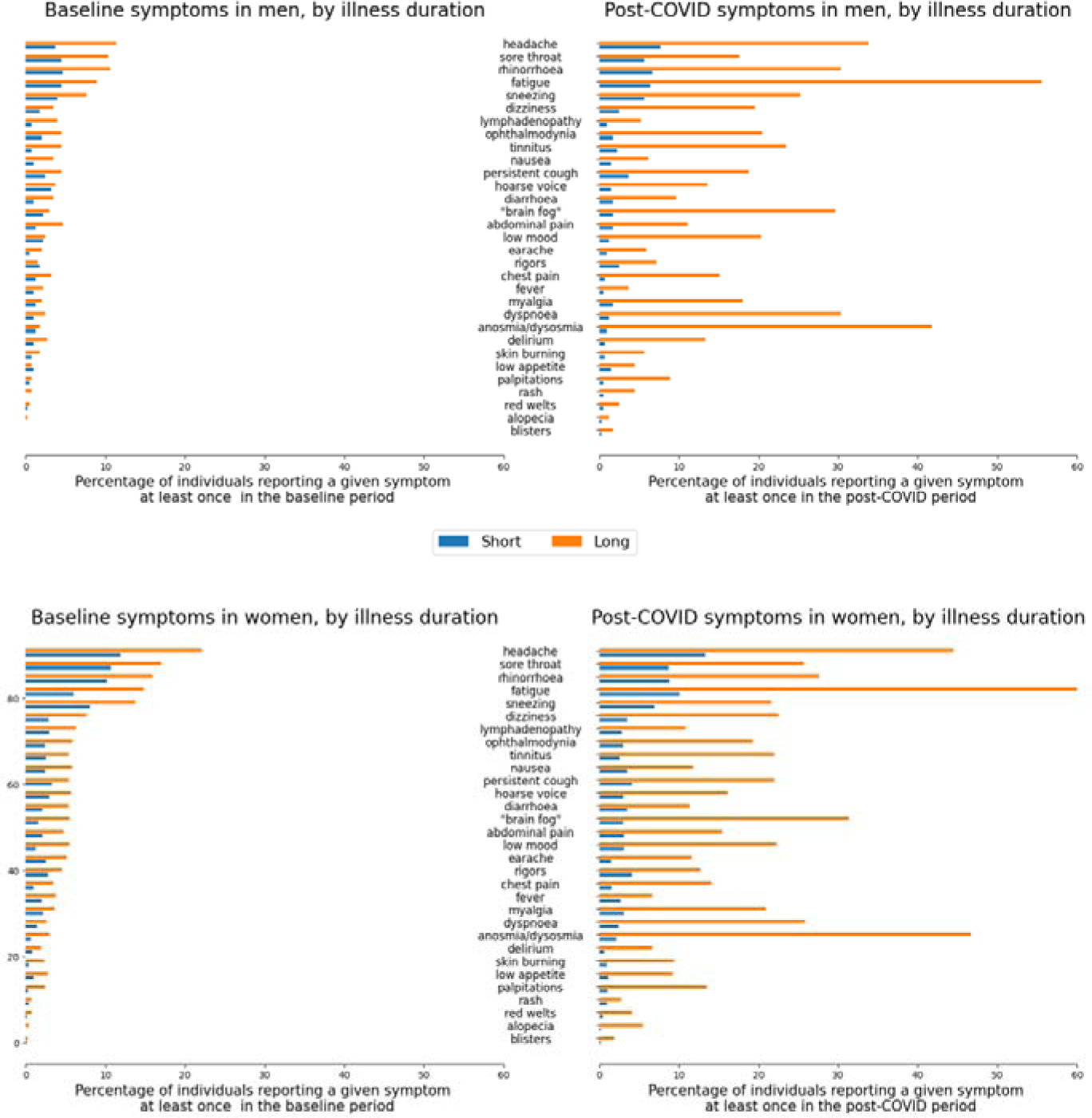
Symptom prevalence by sex, considered during baseline (left two panels) and post-COVID (right two panels) periods, in individuals with short (upper two panels) and long (lower two panels) illness.

### Symptom severity over time

Considering individuals with long illness and fatigue at baseline (176 individuals): 101 (57%) reported unchanged, 45(26%) improved, and 30 (17%) worsened fatigue severity. Considering individuals with long illness with dyspnoea at baseline (35 individuals): 22 (65%) reported unchanged, 11(31%) improved, and 2 (6%) worsened severity (**Suppl. Table S1; Suppl. Fig. S3)**.

For comparison, individuals with short illness duration showed improvement or resolution of either symptom if reported at baseline, noting again the bias imposed by our inclusion criteria.

Considering the 22 individuals with asthma/lung disease reporting dyspnoea at baseline (independent of disease duration): 1 (4.5%) reported worsening, 13 (59%) unchanged; and 8 (36%) decreasing or resolved dyspnoea.

### Baseline symptoms and seasonality

Individuals with long illness were more likely to be asymptomatic at baseline if this period occurred during May-September vs. other times of year (280 of 910 [30.8%] vs. 97 of 440 [22.0%], p=0.002). Reciprocally, the odds ratio for experiencing some specific symptoms at baseline was higher in November-March vs. May-September (for low mood, headache, rhinorrhoea, lymphadenopathy, chest pain, sneezing, sore throat, and ophthalmodynia), after correcting for all Model 3 covariates (data not shown).

Individuals with short illness also appeared to have more rhinorrhea, sneezing, and headache at baseline between November and March, though not significantly after FDR adjustment.

### Baseline symptoms in individuals with long illness

In individuals with long illness, 440 (32.6%) reported at least one symptom at baseline whereas 910 (67.4%) were asymptomatic.

Symptomatic individuals were more likely to be female (336 of 440 [76.4%] vs. 609 of 910 [66.9%], p=0.0004); younger (54 years [46; 62] vs. 59 years [52; 65], p<0.0001); have allergic rhinitis (248 of 440 [56.4%] vs. 404 of 910 [44.3%], p<0.0001) and/or a prior mental health diagnosis (141 of 440 [32.0%] vs. 230 of 910 [25.7%], p=0.011). Thirty-six of 507 (7.1%) symptomatic and 61 of 1088 (5.6%) asymptomatic individuals were health care workers (p=0.29)).

Post-COVID symptom burden was higher in individuals with long illness who were symptomatic (vs. asymptomatic) at baseline (median [IQR] symptom burden (6 [3; 9] vs. 4 [2; 7], p<0.0001). Baseline symptom burden was associated with post-COVID symptom burden (p<0.0001; β=+5.6% CI [4.4; 6.8] per additional baseline symptom) after adjustment for all initial matching criteria.

Baseline symptomatic status (here, any symptom) did not affect the odds of experiencing post-COVID symptoms of cutaneous manifestations, palpitations, dyspnoea, cough, tinnitus, hoarse voice, fever, rigors, anorexia, or dizziness. All other post-COVID symptoms were more common in individuals with long illness and any symptom at baseline, with the exception of anosmia/dysosmia which was less likely post-COVID (OR=0.75 [0.58; 0.96], p=0.022) (Figure 7)

**Figure 7:**
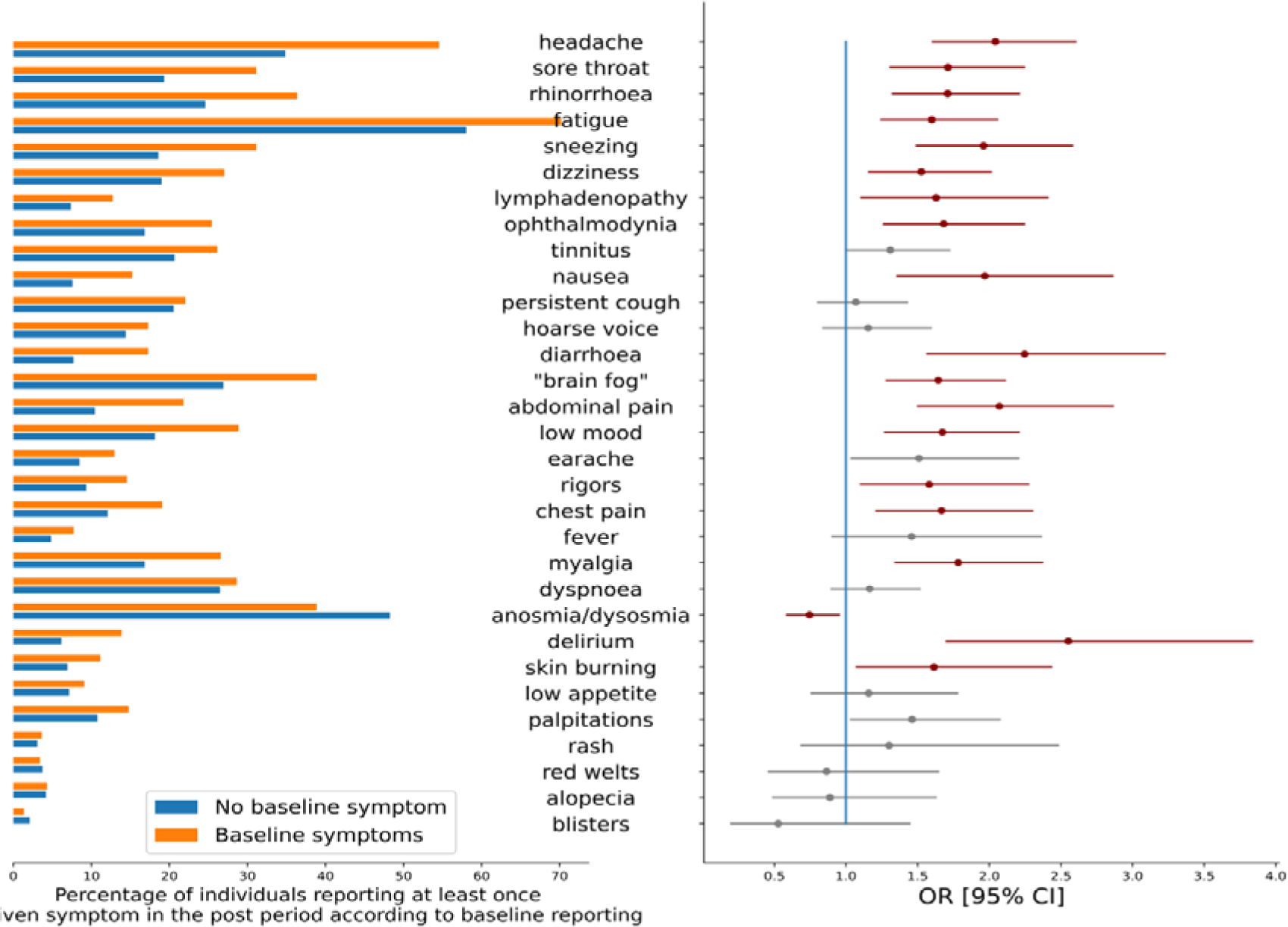
Symptom prevalence in individuals with long illness according to baseline symptom reporting (any symptom reported vs. no symptom); odds ratios of each individual symptom in the post-COVID period according to baseline symptom reporting (any reported symptom vs. no symptom). Red lines indicate significantly increased odds. Symptoms are ordered by decreasing prevalence at baseline.

### Influence of prior comorbidities on baseline and post-COVID symptoms

Having at least one prior comorbidity (here, including prior mental health diagnosis) was more common in individuals with long vs. short illness (926 of 1350 (68.6%) individuals with long illness vs. 668 of 1350 (49.5%) individuals with short illness; p<0.001). Individuals with long illness duration and at least one prior comorbidity (vs. those without) were more likely to have baseline symptoms (here, any symptom) (326 out of 926 [35.2%] vs. 114 out of 424 [26.9%], p=0.003); and experienced greater post-COVID symptom burden (5 [2; 9] vs. 3 [2; 6], p<0.0001) (Figure 8).

**Figure 8.**
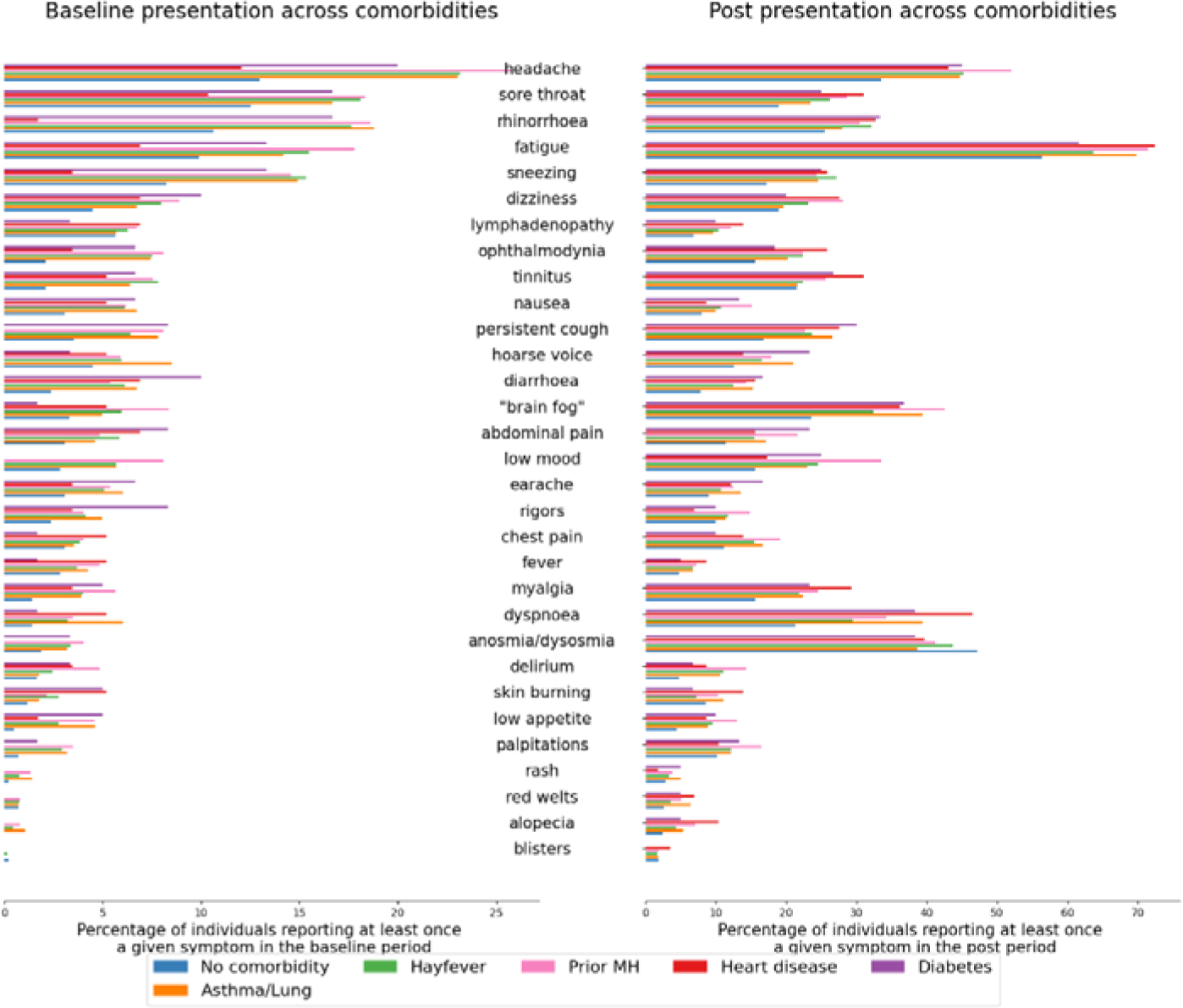
Symptom prevalence in individuals with long illness duration, comparing individuals with respiratory illness (asthma and/or ‘lung disease’, allergic rhinitis), mental health diagnosis, heart disease or diabetes to individuals without any comorbidity, during baseline (left panel) and post-COVID (right panel) periods. MH: prior mental health diagnosis. Symptoms are ordered by decreasing prevalence at baseline.

Odds ratios for individual symptoms experienced during baseline and post-COVID periods in individuals with at least one prior comorbidity, per comorbidity, are shown in **Suppl. Fig S4**.

## Sensitivity analysis

Easing logging regularity to fortnightly-minimum reporting and (consequently) defining end of illness as two weeks of healthy reports, then reselecting individuals for the matched study, gave remarkably stable results. Samples sizes increased to 2496 individuals per group. Prevalence of baseline symptoms in individuals with long illness was similar to previously, and again nearly twice that of individuals with short illness (794 [31.8%] vs 441 [17.7%]). The five commonest symptoms at baseline were unchanged, and again the same in both groups, in slightly different order and proportions (data not shown). As previously, all comorbidities (except kidney disease and cancer) and a prior mental health diagnosis were more common in individuals with long illness. Suppl. Fig S5 presents raw symptom prevalences at baseline for short and long illness groups; Suppl. Figs S6 and S7 display OR for symptom consistency between baseline and follow-up in individuals with short and long duration respectively.

## Discussion

Here we have shown a relationship between symptoms before COVID-19 and subsequent illness duration. Overall, individuals with long illness were nearly twice as likely to report symptoms 4-8 weeks before SARS-CoV-2 infection than individuals with short illness (32.5% vs. 18.0%). However, two-thirds of individuals with long illness were asymptomatic before COVID-19.

The odds of long illness increased for any baseline symptom, and for most individual symptoms. Baseline and post-COVID individual symptoms correlated closely in individuals with long illness, less evident in individuals with short illness, acknowledging the bias created by our selection criteria.

Commonest baseline symptoms, regardless of illness duration, were rhinorrhea, sneezing, sore throat, fatigue and headache, each with higher prevalence in individuals with subsequent long (vs. short) illness. The cause of these baseline non-specific symptoms is unclear, noting here low UK circulation of respiratory viruses beyond SARS-CoV-2 during the time period of this study [19].and that health care workers (with possibly greater workplace exposure to respiratory viruses) did not differ in baseline symptom status. Baseline symptoms might reflect non-infectious comorbidities and individuals with several prior comorbidities (most commonly: allergic rhinitis, asthma/lung disease, and a prior mental health diagnosis) were more likely to report symptoms during baseline and post-COVID periods **F**(**igure 8**) though no clear differential symptom pattern within these groups was evident (**Suppl. Figure S4**).

Despite higher UK pollen counts in May-September, individuals with long illness were less likely to have baseline symptoms during this time. Several symptoms, including low mood, were more common in individuals with long illness and November-March baselines, noting seasonal affective disorder was not solicited in the mental health questionnaire [15].

Our data suggests that some post-COVID symptoms, particularly in individuals with prior comorbidities, may reflect other, serious, non-COVID illness(es). If so, symptom misattribution to OSC/PCS might cause suboptimal management of these other illness(es), with persistence and/or worsening of the other condition consequently. Alternatively, individuals with these comorbidities might be at greater risk of SARS-CoV-2 infection, or of more severe COVID-19 [3]); their underlying comorbidities might be exacerbated by SARS-CoV-2 infection; and/or they may be more vulnerable to specific new pathologies initiated by SARS-CoV-2 infection. For example, post-COVID dyspnoea in individuals with asthma might represent usual asthma, post-viral asthma exacerbation, and/or superimposed pathologies specific to SARS-CoV-2 infection such as post-pneumonitis fibrosis or pulmonary microembolism (pertinently, our data did not support worsening dyspnoea in individuals with asthma/lung disease; and recent systematic reviews and meta-analyses show asthma was associated with lower risk of SARS-CoV-2 infection or of severe COVID-19 [20]. Lastly, altered pandemic health-care access might disproportionately affect individuals with prior comorbidities; however, there was no evidence of such differential access to UK primary care during the pandemic.

Our data concord with two large retrospective studies using primary care data [10,21]. As mentioned, a large UK study of community-managed adults with (n=486,149) and without (n=1,944,580) SARS-CoV-2 infection showed moderately higher symptom prevalences in individuals with (vs. without) SARS-CoV-2 twelve weeks after index event, though this difference narrowed over time [10]. Similar to our analysis, longer symptom duration associated with female sex, younger age, and several prior comorbidities including respiratory illnesses and mental health diagnoses; in contrast to our analysis, this study did not compare pre-vs. post-infection symptoms per-individual [10]. A German study of 51,630 general practice patients with COVID-19 reported PCS prevalence of 8.3% (without comparison population), associated with female sex, comorbidities of asthma and several mental health disorders, and, in contrast to our data, older age; prior symptoms (as opposed to prior co-morbidities) were not assessed [21]. Our data also concord with a study of three longitudinal cohorts (54960 participants, 3193 testing positively for SARS-CoV-2, of whom 1403 developed OSC/PCS), which showed pre-infection psychological distress associated with increased risk of post-COVID symptoms [22]. Pertinently, the UK lockdown abrupt increased mental distress, particularly in females, younger adults, individuals with young children, and individuals with pre-existing mental health conditions [23]; and lockdown had a disproportionate effect on symptom experience in individuals with pre-existing mental health vulnerabilities [24].

Our data also concord with a sub-study of the observational Dutch Lifelines cohort [25], which prospectively collected symptom data across the pandemic. Symptoms were considered pre- and post-infection (out to 90-150 days after illness onset, or matched time point) in 4231 individuals with COVID-19 (both community-and hospital-managed individuals) and 8462 controls. Symptom severity worsened more in the COVID-19 group (vs. uninfected controls), both during acutely and during days 90-150, for several symptoms including breathlessness, chest pain, myalgias, anosmia, and fatigue: overall, 21.4% of cases (vs. 8.7% of controls) had substantial symptom increases 90-150 days after infection compared with pre-infection, an increased symptom severity burden of 12.7% above background. Again, a gender effect was evident: females reported greater symptom severity acutely and longer persistence of increased symptom severity, than males. In contrast, we assessed only individuals with confirmed COVID-19, although our baseline pre-infection data also provide insights into background community symptom prevalences during the pandemic.

Lastly, nocebo effects need consideration. Media commentary regarding OSC/PSC has been widespread, often featuring ‘floating numerators’. Individuals with anxiety and psychological distress are particularly vulnerable to nocebo effects for pain [26]; whether applicable to the pandemic experience is unknown. Relevantly, a high nocebo effect was observed post-SARS-CoV-2 vaccination [27].

## Sex and age effects

In both short and long illness groups, both sexes were similarly assiduous and persistent in reporting (data not shown), contributed to by study design. Nonetheless, whether ultimately experiencing long or short illness, individuals with baseline symptoms were more likely to be female, and younger, than their asymptomatic counterparts. Our results concord with previous studies showing higher symptom reporting by females vs. males for many conditions, including symptom prevalence in daily life [28] and post-acute infection syndromes [11]. Sex differences in illness presentation are increasingly recognised, with potential for differential gender-discriminatory healthcare. Our data also concord with previous studies showing decreased symptom reporting (particularly stress-related symptoms) with age [29].

## Strengths and Limitations

Our study used prospective, dense, and granular symptom reporting by each person in a large cohort over an extended time period, across confirmed SARS-CoV-2 infection, irrespective of ultimate illness duration, with each person serving as their own control. Our design and matching approach limited reporter bias, with no (or minimal) difference in logging frequency, duration, or assiduousness by baseline status or by illness duration. Although predominant circulating SARS-CoV-2 variants altered during this study, with differing risks of long illness across variants [16], we controlled for this by matching by testing week. We have avoided the phrase ‘Long COVID’: we do not have health records access and individuals were not asked about this diagnosis formally. Thus, our data are agnostic to self-identification, in contrast to ONS data that included in their definitions anyone with self-defined long COVID of more than four weeks [8]. Lastly, our data are from individuals with community-managed COVID-19, whereas most papers interrogating long symptom duration post-COVID-19 are dominated by data from hospitalised individuals in whom mechanisms underlying symptom duration are likely to be different.

We acknowledge that we have not captured participants’ changing social circumstances over time which may affect symptom experience (for example, varying personal/regional lockdown requirements resulting in varying exposure to other (i.e., non SARS-Cov-2) viruses [19]; varying extent and duration of social isolation and loneliness; differential home-schooling responsibilities which burden was disproportionally experienced by women and associated with markedly increased psychological distress [15]

The requirement for one week’s asymptomatic logging record prior to COVID-19 symptom onset (to enable illness duration to be calculated [3]) may have biased our sample towards healthier people less affected by pre-existing co-morbidities, resulting in underestimation of the relationship of prior symptoms and comorbidities with subsequent illness duration. Additionally, all individuals had community-managed COVID-19; our data cannot be extrapolated to hospitalised individuals.

Although the direct symptom questions expansion from November 2020 was informed by feedback from individuals experiencing OSC/PCS, pertinent symptoms may have been missed - although our direct questions covered the symptom groups in other PCS studies [10,21,25]. Further, all but two symptom questions were binary (yes/no) with no available quantitative assessment or health record linkage.

Our analysis used data from the first reported SARS-CoV-2 positive result and subsequent illness duration. Evidence of repeated infection was used as demographic matching criteria (45 in each group); however, as it transpired, only one illness episode was included per participant.

Overall, ZOE/CSS app users are not representative of the UK population (younger, more female, higher educational status, lower ethnic diversity, over-representative of healthcare workers). Moreover, individuals had to log assiduously, at length, which might bias our data towards individuals with specific app usage behaviours and potentially exclude individuals with app usage fatigue. To assess this we compared our study participants to all ZOE app symptomatic test-positive users, across the same time period: our cohort were more persistent than positive ZOE app users overall (time between symptom commencement and last report: median 206 days [IQR: 140;292] for our cohort vs. 99 days [IQR: 15;159] for test-positive symptomatic ZOE app users overall); and app fatigue was not evident but the opposite, with strong correlation (Spearman rho 0.465, p<0.00001) in all symptomatic test-positive app users between symptom duration and continuing app usage (both calculated from date of symptom onset). Although we cannot comment more granularly (e.g., possible differential drop-out of individuals with particular symptoms) these data do not support app fatigue; and pertinently, two-thirds of our long illness group had symptoms for ≥12 weeks.

Our inclusion criteria required participation in the mental health questionnaire in early 2021 [15]. Of 1,257,278 million app users at the time, 715,324 (56.8%) participated; whether participation was affected by presence/absence of a prior mental health diagnoses cannot be determined. However, censoring for mental health survey participation only significantly affected the male/female ratio (females more likely to participate; data not shown).

We considered the impact of stringent logging frequency possibly precluding individuals with longer and/or more severe illness. However, our sensitivity analysis with loosened stringency did not change our results significantly (**Supp. Fig. S5-S7**).

Lastly, we cannot exclude an effect of app participation *per se* on symptom reporting (regardless of disease duration), noting that use of symptom tracking apps can inflate symptom reporting in some individuals [30].

## Conclusion

Symptoms prevalence before SARS-CoV-2 infection differed in individuals who subsequently manifest long vs. short illness, with correlation of symptom burden and specific symptom experience between baseline and post-COVID periods. At least some of this risk is influenced by prior comorbidities. However, the majority of individuals with post-COVID symptoms were asymptomatic before SARS-CoV-2 infection. Our data highlight the importance of parsing symptoms related to COVID-19 vs. other diseases – but also the difficulties of this. Genetic studies in OSC/PCS, including Mendelian randomization approaches, may prove unbiased means of disentangling association from causation. Practically speaking, the clinical implications are unchanged: an open and holistic approach is needed to manage post-COVID symptoms, whatever their aetiology.

## Data Availability

Data collected in the COVID Symptom Study smartphone app can be shared with other health researchers through the UK National Health Service-funded Health Data Research UK and Secure Anonymised Information Linkage consortium, housed in the UK Secure Research Platform (Swansea, UK). Anonymised data are available to be shared with researchers according to their protocols in the public interest

https://web.www.healthdatagateway.org/dataset/594cfe55-96e3-45ff-874c-2c0006eeb881.

## Acknowledgements

We acknowledge with gratitude the dedication and commitment of the contributors of the Zoe app to citizen science enabling this research.

## Data sharing statement

Data collected in the COVID Symptom Study smartphone app can be shared with other health researchers through the UK National Health Service-funded Health Data Research UK and Secure Anonymised Information Linkage consortium, housed in the UK Secure Research Platform (Swansea, UK). Anonymised data are available to be shared with researchers according to their protocols in the public interest https://web.www.healthdatagateway.org/dataset/594cfe55-96e3-45ff-874c-2c0006eeb881.

## Contribution statement

ELD and CHS conceptualised the study design. MA, BM, JCP, CHS contributed to data curation and associated software tools. CJS, TS contributed to acquisition design. CHS performed the analysis and verified the underlying data. CHS interpreted the results with ELD who supervised the study. ELD and CHS drafted the manuscript. TDS, SO and CJS acquired funding. All authors contributed to interpretation of data and critical revision of the manuscript.

## Transparency statement

The lead author affirms that the manuscript is an honest, accurate, and transparent account of the study being reported; that no important aspects of the study have been omitted; and that any discrepancies from the study as planned have been explained.

