## Supplemental material for "Symptom experience before vs. after confirmed SARS-CoV-2 infection: a population and case control study using prospectively recorded symptom data"

**Supplementary Tables and Figures**

**Supplementary Table S1**: **List of symptom and comorbidity questions asked in the COVID Symptom Study application during the current study period.**

| **Description on the app** | **Symptom** |
| --- | --- |
| Fever (at least 37.8 C or 100F) | Fever |
| Persistent cough (coughing a lot for more than an hour or 3 or more coughing episodes in 24 h) | Cough |
| Unusual fatigue:   - No - Mild - Severe fatigue – I struggle to get out of bed | Fatigue |
| Shortness of breath or trouble breathing   - No - Yes. Mild symptoms – slight shortness of breath during ordinary activity - Yes. Significant symptoms – breathing is comfortable only at rest - Yes. Severe symptoms – breathing is difficult even at rest | Dyspnoea |
| Loss of smell / taste | Anosmia |
| Unusually hoarse voice | Hoarse voice |
| Unusual chest pain or tightness in your chest | Chest pain |
| Unusual abdominal pain or stomachache | Abdominal pain |
| Diarrhoea | Diarrhoea |
| Confusion, disorientation, drowsiness | Delirium |
| Skipping meals | Low appetite |
| Headache | Headache |
| Nausea or vomiting | Nausea |
| Dizziness or light-headedness | Dizziness |
| Unusual eye soreness or discomfort (light sensitivity, excessive tears, pink/red eye) | Ophthalmodynia |
| Sore or painful throat | Sore throat |
| Unusual strong muscle pains or aches | Myalgia |
| Raised red itchy welts on skin or sudden swelling of the face or lips | Red welts |
| Red/purple sores or blisters on your feet including your toes | Blisters |
| Rash on your arms or torso | Rash |
| Strange unpleasant sensations in your skin like pins and needles or burning | Skin burning |
| Unusual hair loss | Alopecia |
| Feeling down, depressed or hopeless | Low mood |
| Loss of concentration or memory (brain fog) | Brain fog |
| Altered smell / taste (things smell or taste different than usual) | Dysosmia |
| Runny nose | Rhinorrhea |
| Sneezing more than usual | Sneezing |
| Earache | Earache |
| Ringing in your ears | Tinnitus |
| Swollen neck glands | Lymphadenopathy |
| Unusually fast or irregular heartbeat (palpitations) | Palpitations |

**Supplementary Table S2:** List of questions regarding prior Mental Health diagnoses, asked as part of a mental health questionnaire between 23 February and 12 April 2021 (33).

Have you ever been diagnosed with a mental health condition?
- Yes
 - No
 - Prefer Not to Say

If the above is Yes:

- Generalised anxiety order (GAD)

- Panic disorder

- Specific phobias

- Obsessive compulsive disorder (OCD)

- Post-traumatic stress disorder (PTSD)

- Social anxiety disorder

- Agoraphobia

- Depression

- Attention deficit or attention deficit and hyperactivity disorder (ADD / ADHD)

- Autism, Asperger's or autistic spectrum disorder

- Eating disorder (e.g., bulimia nervosa; anorexia nervosa; psychological over-eating or binge-eating)

- A personality disorder

- Mania, hypomania, bipolar or manic depression

- Schizophrenia

- Substance use disorder

- Any other type of psychosis or psychotic illness

- Prefer not to say

- Other

**Supplementary Table S3: Symptoms during the baseline period.** Symptoms are ordered by prevalence in the group with long illness duration at baseline.

| **Symptom reported during post-COVID period (irrespective of subsequent illness duration)** | **Female (n) Total=1890** | **Female (%)** | **Male (n) Total=810** | **Male (%)** | **p-value** |
| --- | --- | --- | --- | --- | --- |
| Headache | 321 | 16.98 | 61 | 7.53 | 0.000 |
| Sore throat | 262 | 13.86 | 60 | 7.41 | 0.000 |
| Rhinorrhoea | 246 | 13.02 | 62 | 7.65 | 0.000 |
| Fatigue | 197 | 10.42 | 54 | 6.67 | 0.003 |
| Sneezing | 206 | 10.90 | 47 | 5.80 | 0.000 |
| Dizziness | 99 | 5.24 | 21 | 2.59 | 0.003 |
| Lymphadenopathy | 87 | 4.60 | 19 | 2.35 | 0.008 |
| Ophthalmodynia | 78 | 4.13 | 26 | 3.21 | 0.305 |
| Tinnitus | 75 | 3.97 | 21 | 2.59 | 0.098 |
| Nausea | 78 | 4.13 | 18 | 2.22 | 0.019 |
| Cough | 82 | 4.34 | 28 | 3.46 | 0.339 |
| Hoarse voice | 82 | 4.34 | 28 | 3.46 | 0.339 |
| Diarrhoea | 71 | 3.76 | 18 | 2.22 | 0.054 |
| ‘Brain fog’ | 67 | 3.54 | 21 | 2.59 | 0.247 |
| Abdominal pain | 65 | 3.44 | 24 | 2.96 | 0.605 |
| ‘Feeling down’ | 64 | 3.39 | 19 | 2.35 | 0.189 |
| Earache | 73 | 3.86 | 10 | 1.23 | 0.000 |
| Rigors | 69 | 3.65 | 13 | 1.60 | 0.007 |
| Chest pain | 42 | 2.22 | 18 | 2.22 | 1.000 |
| Fever | 55 | 2.91 | 13 | 1.60 | 0.064 |
| Myalgia | 55 | 2.91 | 13 | 1.60 | 0.064 |
| Dyspnoea | 38 | 2.01 | 14 | 1.73 | 0.737 |
| Anosmia/Dysosmia | 34 | 1.80 | 12 | 1.48 | 0.673 |
| Delirium | 26 | 1.38 | 15 | 1.85 | 0.450 |
| Skin burning | 26 | 1.38 | 10 | 1.23 | 0.913 |
| Anorexia | 35 | 1.85 | 7 | 0.86 | 0.083 |
| Palpitations | 26 | 1.38 | 5 | 0.62 | 0.134 |
| Rash | 11 | 0.58 | 3 | 0.37 | 0.682 |
| Red Welts | 8 | 0.42 | 3 | 0.37 | 1.000 |
| Alopecia | 4 | 0.21 | 1 | 0.12 | 1.000 |
| Blisters | 3 | 0.16 | 0 | 0.00 | NA |

**Supplementary Table S4: Symptom prevalence during the post-COVID period, irrespective of illness duration.** Symptoms are ordered by prevalence in the group with long illness duration at baseline.

| **Symptom reported during post-COVID period (irrespective of subsequent illness duration)** | **Females (n)** | **Females (%)** | **Males (n)** | **Males (%)** | **p-value** |
| --- | --- | --- | --- | --- | --- |
| Headache | 546 | 28.89 | 168 | 20.74 | 0.000 |
| Sore throat | 324 | 17.14 | 94 | 11.60 | 0.000 |
| Rhinorrhoea | 344 | 18.20 | 150 | 18.52 | 0.888 |
| Fatigue | 707 | 37.41 | 251 | 30.99 | 0.002 |
| Sneezing | 269 | 14.23 | 125 | 15.43 | 0.454 |
| Dizziness | 246 | 13.02 | 89 | 10.99 | 0.161 |
| Lymphadenopathy | 129 | 6.83 | 25 | 3.09 | 0.000 |
| Ophthalmodynia | 210 | 11.11 | 90 | 11.11 | 1.000 |
| Tinnitus | 232 | 12.28 | 104 | 12.84 | 0.731 |
| Nausea | 144 | 7.62 | 31 | 3.83 | 0.000 |
| Cough | 247 | 13.07 | 91 | 11.23 | 0.209 |
| Hoarse voice | 180 | 9.52 | 61 | 7.53 | 0.112 |
| Diarrhoea | 140 | 7.41 | 46 | 5.68 | 0.123 |
| ‘Brain fog’ | 324 | 17.14 | 127 | 15.68 | 0.380 |
| Abdominal pain | 175 | 9.26 | 52 | 6.42 | 0.018 |
| ‘Feeling down’ | 239 | 12.65 | 87 | 10.74 | 0.184 |
| Earache | 124 | 6.56 | 28 | 3.46 | 0.002 |
| Rigors | 159 | 8.41 | 39 | 4.81 | 0.001 |
| Chest pain | 148 | 7.83 | 64 | 7.90 | 1.000 |
| Fever | 88 | 4.66 | 17 | 2.10 | 0.002 |
| Myalgia | 226 | 11.96 | 80 | 9.88 | 0.134 |
| Dyspnoea | 267 | 14.13 | 128 | 15.80 | 0.285 |
| Anosmia/Dysosmia | 461 | 24.39 | 173 | 21.36 | 0.098 |
| Delirium | 69 | 3.65 | 57 | 7.04 | 0.000 |
| Skin burning | 98 | 5.19 | 26 | 3.21 | 0.032 |
| Anorexia | 98 | 5.19 | 24 | 2.96 | 0.014 |
| Palpitations | 137 | 7.25 | 38 | 4.69 | 0.017 |
| Rash | 35 | 1.85 | 20 | 2.47 | 0.372 |
| Red welts | 43 | 2.28 | 12 | 1.48 | 0.234 |
| Alopecia | 54 | 2.86 | 6 | 0.74 | 0.001 |
| Blisters | 20 | 1.06 | 8 | 0.99 | 1.000 |

**Supplementary Figure S1**

**Symptom prevalence by duration, considered during baseline (left panels) and post-COVID (right panels) periods, in males (upper panels) and females (lower panels).**

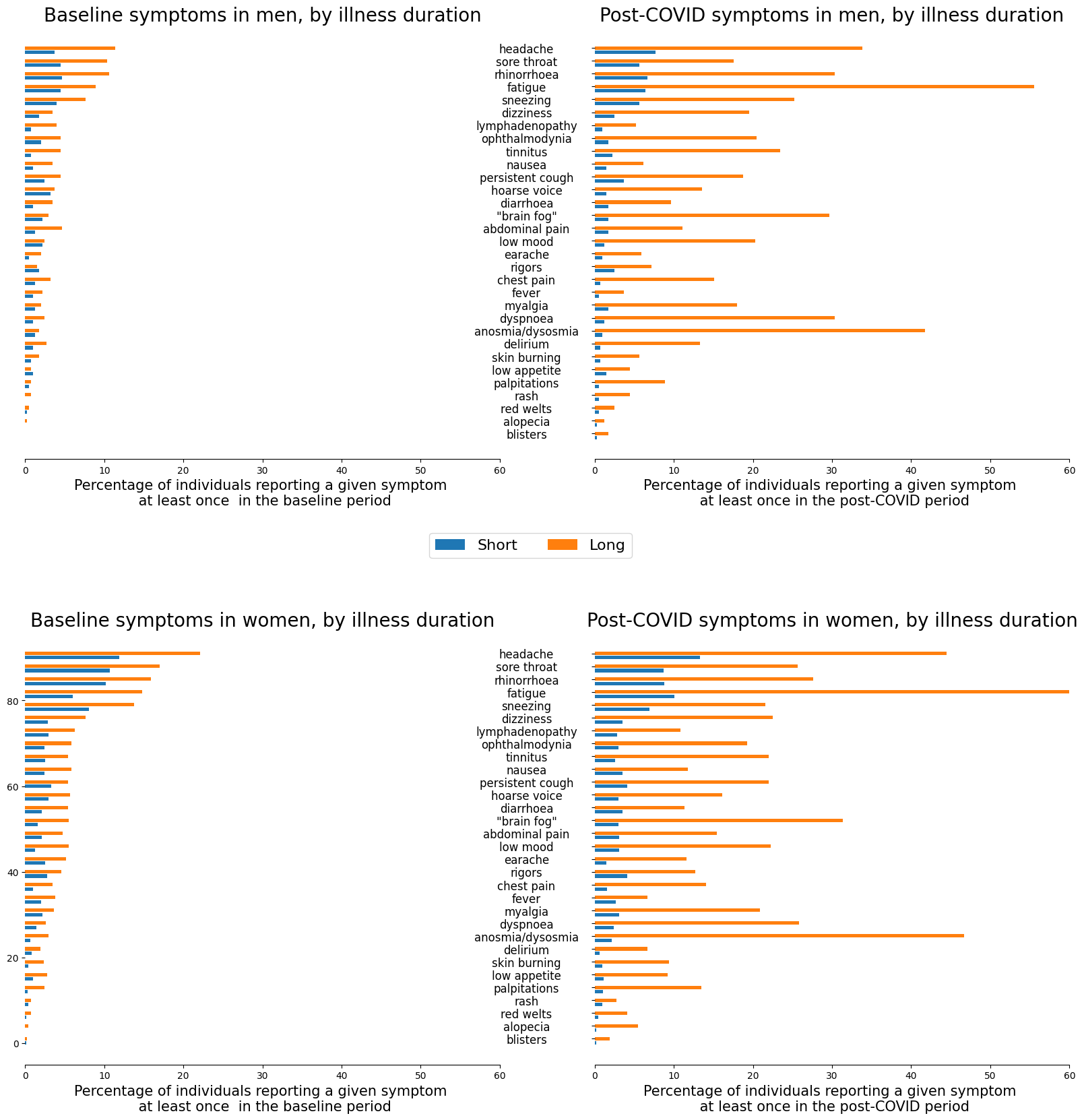

**Supplementary Figure S2 Odds ratios of symptom concordance (i.e., present in the post-COVID period, if reported during the baseline period) in individuals with short illness duration.** Model 1: adjusted for age, sex, BMI, vaccination number, week of testing, smoking, and IMD; Model 2: additionally adjusted for co-morbidities reported at registration; Model 3: additionally adjusted for prior mental health diagnosis. Circle size refers to symptom prevalence during baseline (gold) and post-COVID (purple) periods (scale shown at bottom of image). Symptoms are ordered by decreasing prevalence during the baseline period in individuals with long illness duration. Odds ratios are shown as (dots) with CI (lines); results in red are significant after correction for multiple comparisons.

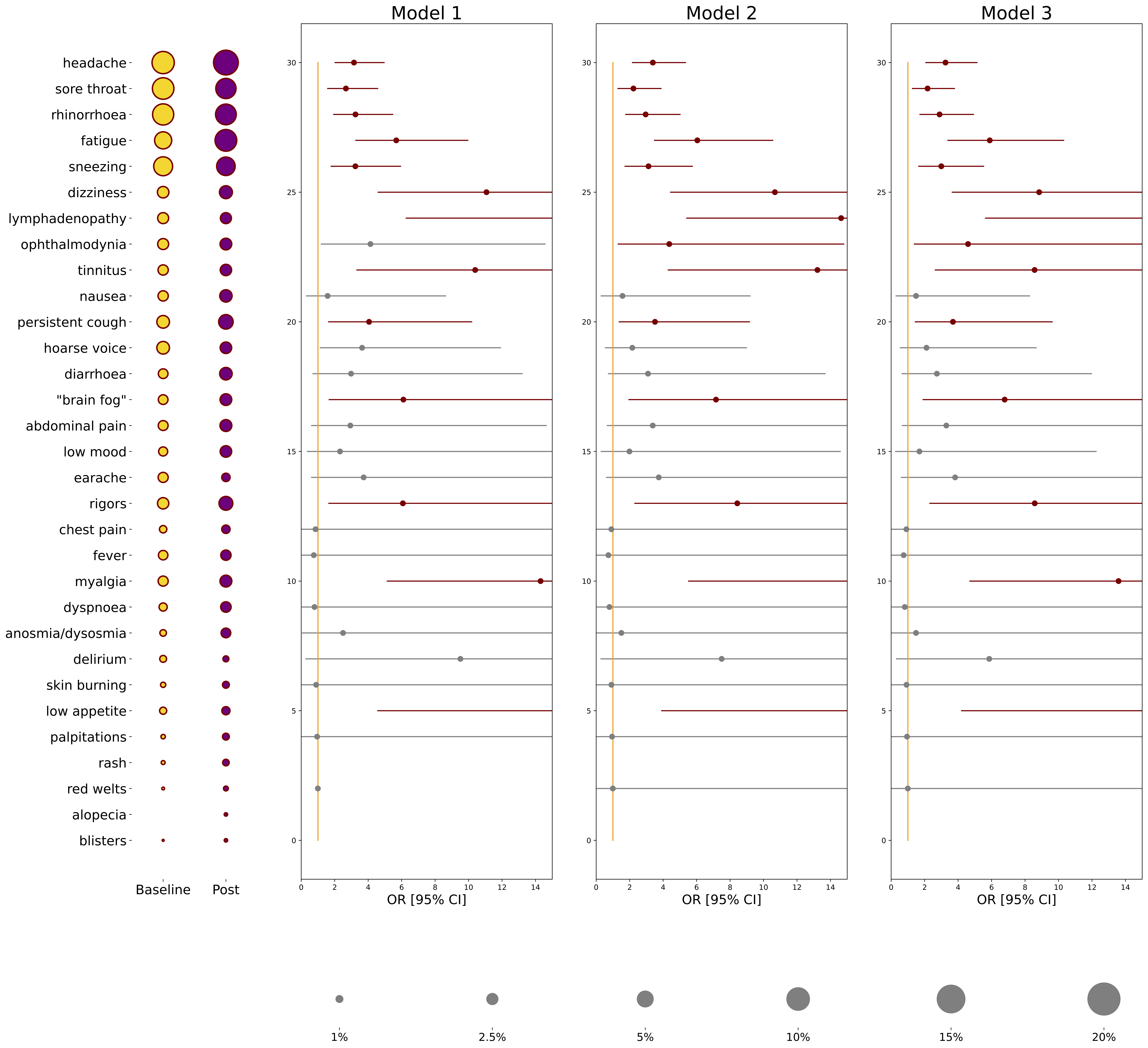

**Supplementary Figure S3: Evolution of Severity of Fatigue and Dyspnoea, in Individuals with Short vs. Long Illness Duration**

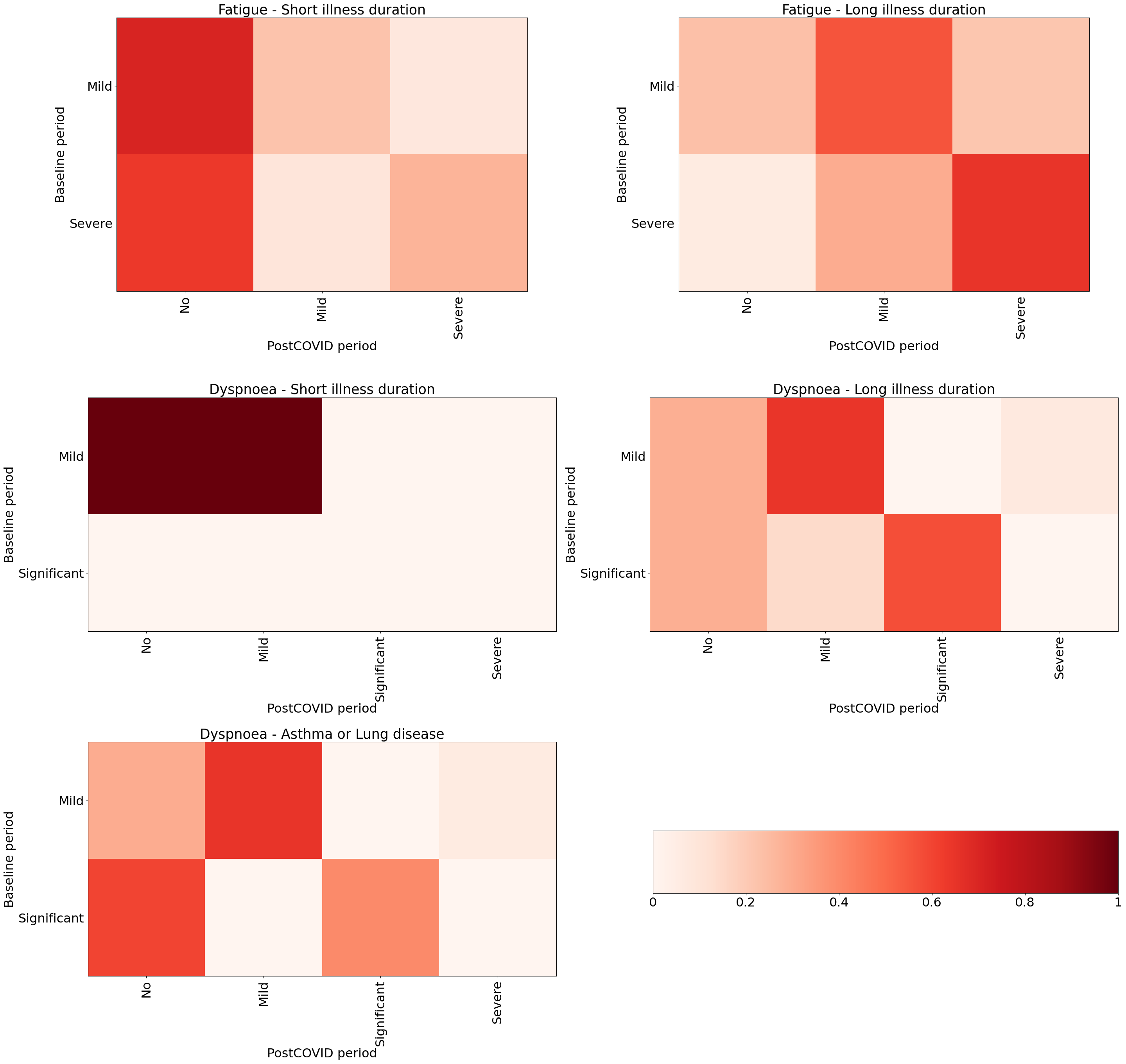

**Supplementary Figure S4:** Odds ratios (colour-coded) for experiencing an individual symptom during baseline (left) and post-COVID (right) periods, for individuals with prior diagnoses of asthma/lung disease, hayfever, a mental health (MH) disorder, heart disease or diabetes. **Symptoms are ordered by prevalence at baseline in individuals with long illness duration.**

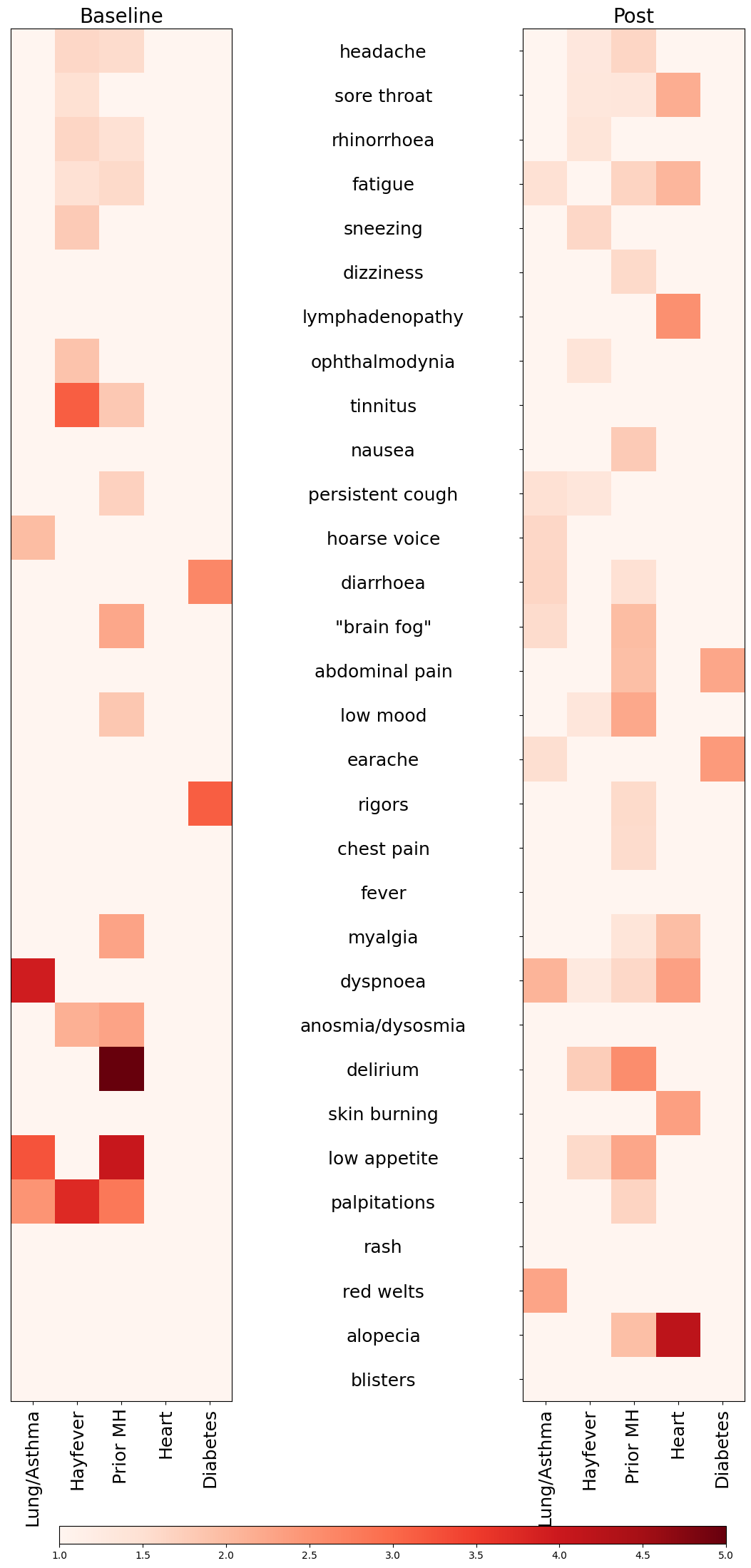

Supplementary Figure S5 - **Symptom prevalence during the baseline period in individuals with long illness vs. short illness when relaxing the logging criteria to every two weeks** (descriptive data only, unadjusted for comorbidities, week of testing, prior infection, vaccination status, smoking or index of multiple deprivation).

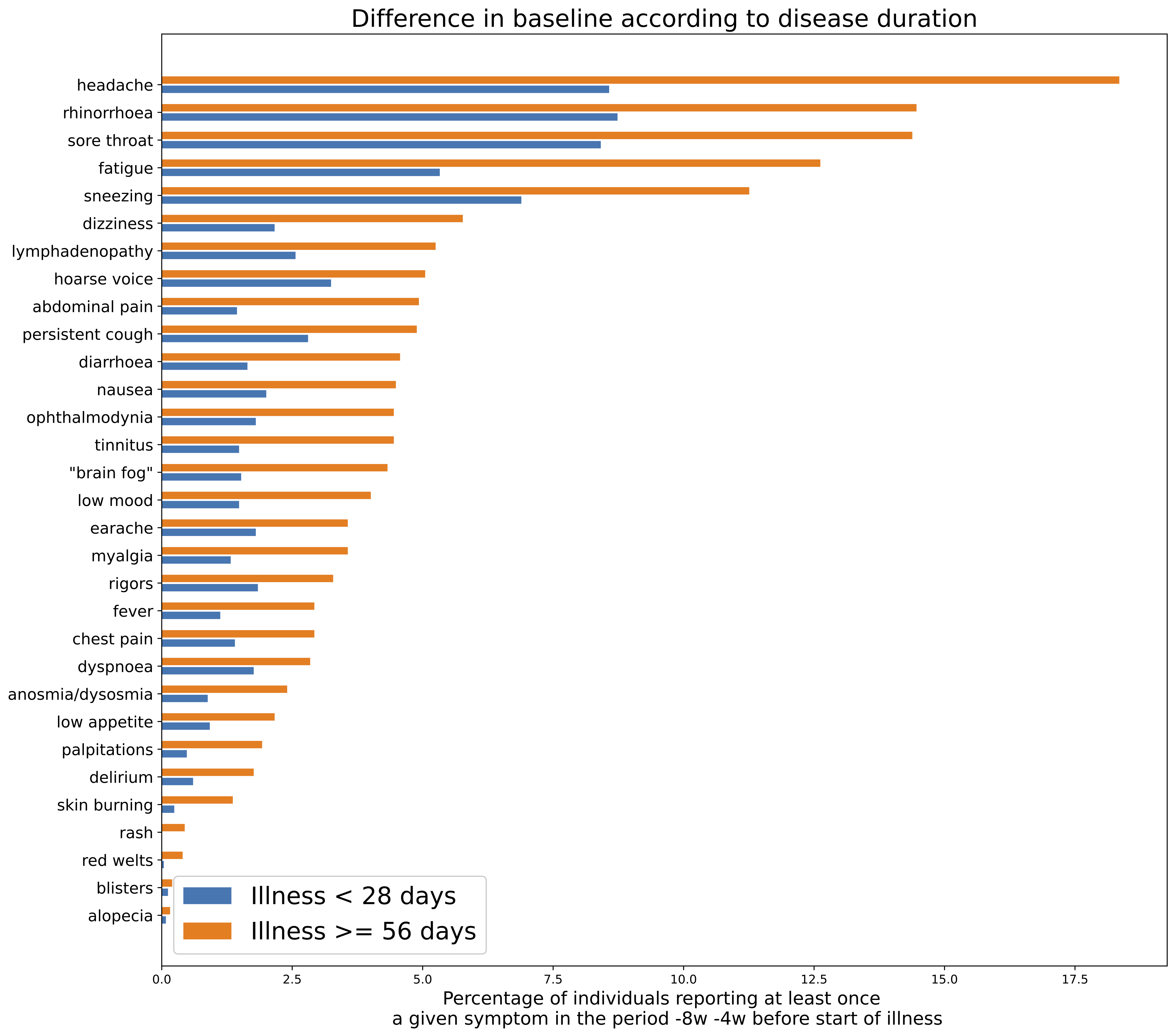

Supplementary Figure S6: **Odds ratios of symptom concordance (i.e., present in the post-COVID period, if reported at baseline [reference period]) in individuals with short illness when relaxing the logging criteria to every two weeks.** Model 1: adjusted for age, sex, BMI, vaccination number, prior infection, week of testing, smoking and index of multiple deprivation; Model 2: additionally adjusted for comorbidities reported at registration; Model 3: additionally adjusted for prior mental health diagnosis. Circle size refers to symptom prevalence during baseline (gold) and post-COVID (purple) periods; scale is shown at bottom of figure. Symptoms are ordered by decreasing prevalence during the baseline period. Odds ratios are shown as dots with CI (lines); results in red are significant after adjustment for multiple comparisons.

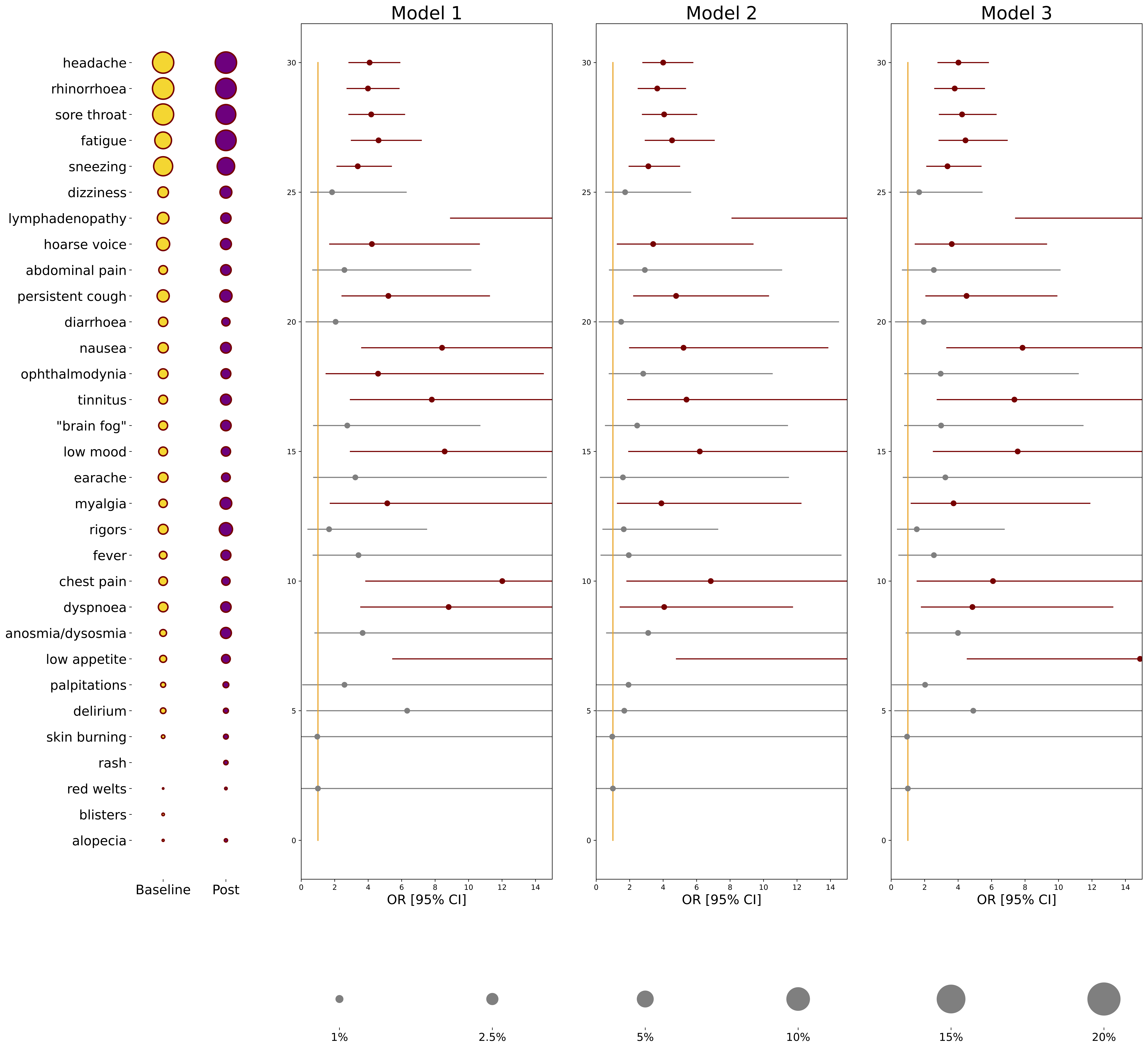

Supplementary Figure S7: **Odds ratios of symptom concordance (i.e., present in the post-COVID period, if reported at baseline [reference period]) in individuals with long illness when relaxing the logging criteria to every two weeks.** Model 1: adjusted for age, sex, BMI, vaccination number, prior infection, week of testing, smoking and index of multiple deprivation; Model 2: additionally adjusted for comorbidities reported at registration; Model 3: additionally adjusted for prior mental health diagnosis. Circle size refers to symptom prevalence during baseline (gold) and post-COVID (purple) periods; scale is shown at bottom of figure. Symptoms are ordered by decreasing prevalence during the baseline period. Odds ratios are shown as dots with CI (lines); results in red are significant after adjustment for multiple comparisons.

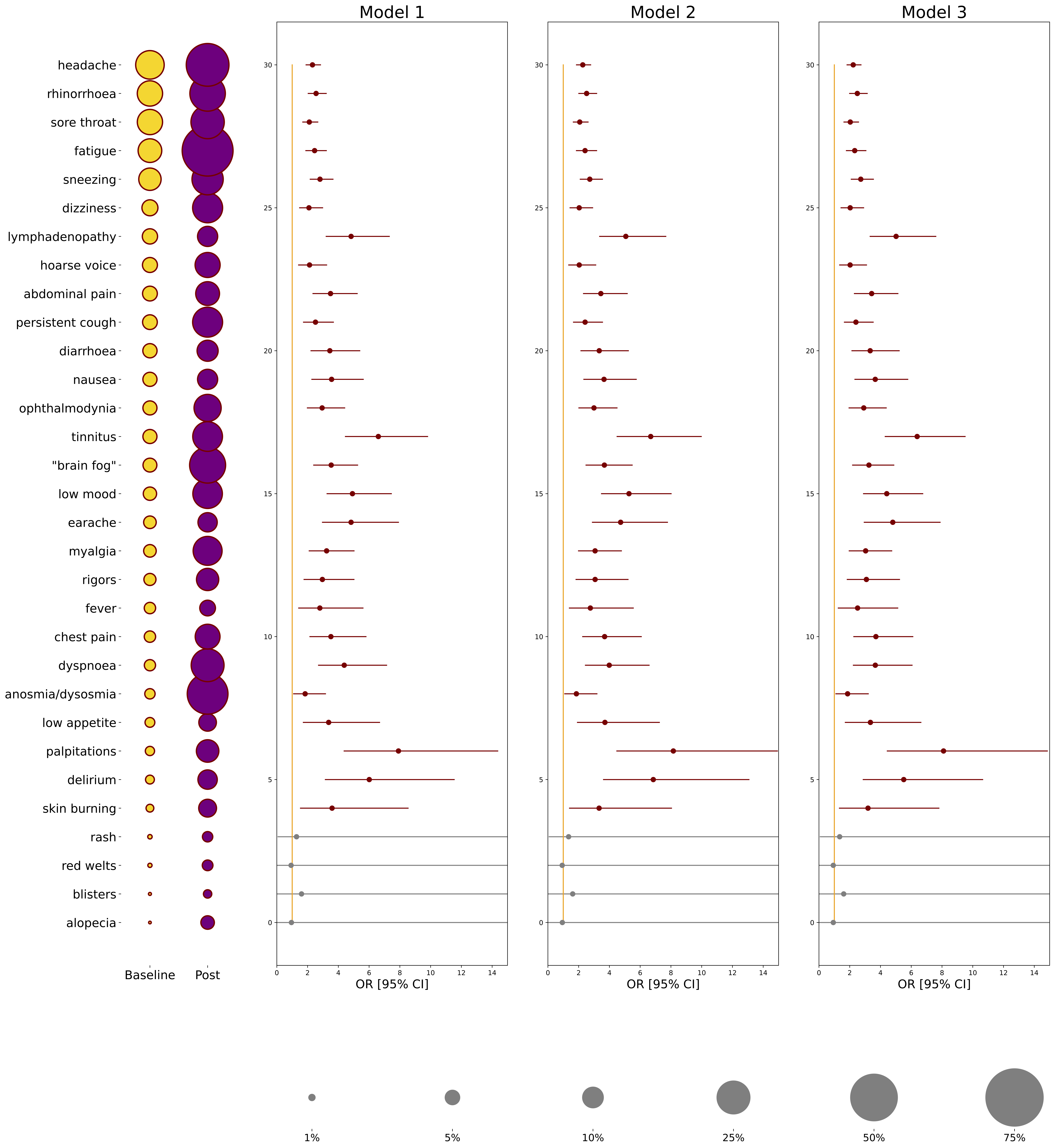
